# Isolating transdiagnostic effects reveals specific genetic profiles in psychiatric disorders

**DOI:** 10.1101/2023.12.20.23300292

**Authors:** Engin Keser, Wangjingyi Liao, Andrea G. Allegrini, Kaili Rimfeld, Thalia C. Eley, Robert Plomin, Margherita Malanchini

## Abstract

Evidence indicates a great degree of genetic overlap between psychiatric diagnoses. Accounting for these transdiagnostic effects can sharpen research on disorder-specific genetic architecture. Here we isolate genetic effects that are shared across 11 major psychiatric disorders (p factor) to gain further insight into genetic specificity and comorbidity over and above that contributed by the p factor, unique to each psychiatric disorder. After adjusting for transdiagnostic genetic effects, we examined genetic correlations among psychiatric traits as well as relationships with other biobehavioural traits. The landscape of genetic associations between pairs of psychiatric disorders changed substantially, and their genetic correlations with biobehavioural traits showed greater specificity. Isolating transdiagnostic genetic effects across major psychiatric disorders provides a nuanced understanding of disorder-specific genetic architecture and genetic comorbidity, and may help guide diagnostic nomenclature and treatment research.

## Introduction

Genetic studies have challenged the current classification of psychiatric disorders as distinct categorical diagnoses by revealing overlaps in their genetic architectures ^1^. For instance, the first genome-wide association study (GWAS) of schizophrenia uncovered shared genetic loci between schizophrenia and bipolar disorder ^2^ despite their distinct categorization in diagnostic manuals (e.g., DSM-IV ^3^). Further research using Linkage Disequilibrium Score Regression (LDSC ^4^) highlighted significant genetic correlations across most psychiatric diagnoses ^5–8^, unlike other neurological disorders such as Parkinson’s and Alzheimer’s disease, which remained genetically distinct ^9^. A recent LDSC analysis of 11 major psychiatric diagnoses found that the genetic correlation between schizophrenia and bipolar disorder was 0.68, and the average of 55 genetic correlations between 11 diagnoses was 0.28 ^10^.

The positive genetic manifold between diagnoses is consistent with the idea of a general factor of psychopathology ^11^, called *p* ^12^. The p factor describes the propensity to developing all forms of psychopathologies ^13^, and reflects the comorbidities between psychiatric conditions that have been observed concurrently ^14^, across the lifespan ^15, 16^, and even across generations ^13, 17^. A p factor has also emerged from recent genetic and genomic studies ^8, 10, 18^. Shared genetic effects across different disorder dimensions were found to be stable over development, even when considering different measures and reporters ^18^. Capturing what cuts across diagnostic categories (i.e., a transdiagnostic approach) was found to be more effective in predicting functional and life outcomes than individual diagnoses ^19^.

Therefore, isolating p from major psychiatric disorders could better capture the specific genetic effects associated with each disorder and provide a more precise understanding of disorder-specific biology. Previous research that isolated transdiagnostic effects to investigate specificity revealed novel genetic profiles and biological pathways in neurodevelopmental disorders ^20^ and alcohol use disorder ^21^. In this study, we applied Genomic Structural Equation Modelling (Genomic SEM) ^22^ to isolate transdiagnostic genetic effects across 11 psychiatric disorders from genetic effects specific to each psychiatric condition. As shorthand, we refer to these disorder-specific genetic effects as *non-p* to describe residual genetic variance independent of genomically identified p. We used summary statistics from these non-p GWA analyses to estimate SNP heritability, genetic correlations between the disorders beyond transdiagnostic effects, and their correlations with external biobehavioural traits. We hypothesized that non-p traits will provide us with novel insight into the genetic architecture of psychiatric disorders and their correlations with other disorders and traits, consequently informing research and practice into diagnostics and treatment.

## Methods

The article is accompanied by <u>Supplementary Information</u> and the study followed a <u>preregistered analysis plan</u>. Deviations from preregistered analyses are described in Supplementary Note 1.

### GWAS summary statistics

We used the most recent publicly available summary statistics from GWA studies of 11 major psychiatric disorders. Detailed information about the GWAS summary statistics, sample sizes and availability are provided in Supplementary Table 1. All summary statistics are based on samples of European ancestry GWAS only. For details on the analysis protocol, we refer to the original publications.

### Genomic SEM

Genomic SEM ^22^ is a statistical framework that can model the shared and unique genetic architecture of complex traits by applying structural equation modelling principles to GWAS summary statistics. Genomic SEM is unbiased by sample overlap and imbalanced sample sizes ^22^. Here, we used Genomic SEM to perform multivariate GWAS analysis of 11 major psychiatric disorders (Figure 1) in order to capture transdiagnostic effects across all 11 disorders and isolate genetic variance in each psychiatric disorder beyond that captured by the p factor.

**Figure 1.**
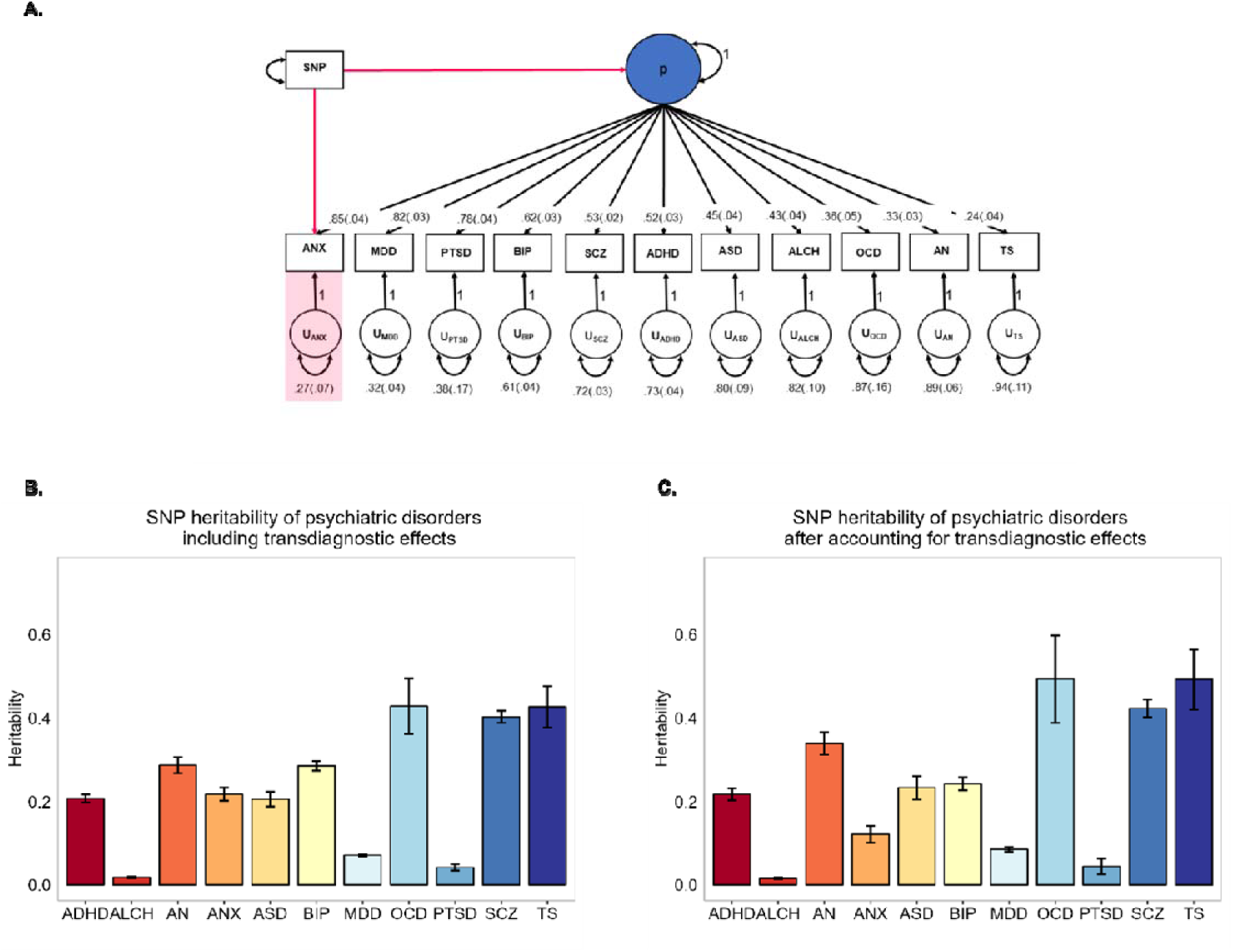
Isolating transdiagnostic genetic effects from 11 major psychiatric disorders. **A.** *Standardized results for a common factor model of genomic p*. Each square indicates observed variables (i.e., the summary statistics for each of the 11 major psychiatric disorders) and circles represent latent variables that are statistically inferred from the data (i.e., genomic p-factor). One-headed arrows are standardized factor loadings, representing regression relations with the arrow pointing from the predictor variable to the outcome variable. Covariance relationships between variables are represented as two-headed arrows linking the variables. Residual variances of a variable are represented as a two-headed arrow connecting the variable to itself. SEs are shown in parentheses. The red arrows linking the SNP to both the p-factor and PTSD provide an example of the model used to partition genetic variance associated with transdiagnostic effects from the genetic variance specific to each disorder. We ran the model 11 times, isolating transdiagnostic effects from each psychiatric condition at a time. ANX = anxiety disorder; MDD = major depressive disorder; PTSD = post-traumatic stress disorder; BIP = bipolar disorder; SCZ = schizophrenia; ADHD = attention-deficit hyperactivity disorder; ASD = autism spectrum disorder; ALCH = problematic alcohol use; OCD = obsessive-compulsive disorder; AN = anorexia nervosa; TS = Tourette syndrome; p = general psychopathology factor. **B and C.** *SNP-based heritability estimates including and after accounting for transdiagnostic effects.* SNP heritabilities were calculated before (on the liability scale) and after (on the observed scale) removing genetic effects shared with the p-factor. Error bars indicate standed errors.

Genomic SEM uses multivariable LD score regression to estimate the genetic covariance matrix and sampling covariance matrix. We applied quality control filters for this step using the defaults in Genomic SEM, including restricting SNPs to those present in HapMap3 with a minor allele frequency > 1% and information score > 0.9. The LD weights used for LDSC were calculated using the European subsample of the 1000 Genomes phase 3 project; excluding the major histocompatibility complex (MHC) due to complex LD structures in this region that can bias estimates. When calculating the liability scale heritability estimates for the uncorrected psychiatric disorders, we used the sum of effective sample sizes, and a sample prevalence of 0.5 to reflect that the corrected sample size already accounts for sample ascertainment. See Supplementary Notes for more details about processing summary statistics in Genomic SEM and calculating effective sample sizes. After the quality control steps, 3,746,806 SNPs were present across all 11 disorders.

### Independent SNPs, Genes, and Enrichment and Pathway Analysis (MAGMA)

To identify independent hits from the 11 non-p GWAS, we applied a pruning approach using a window of 250 kb and an linkage disequilibrium (LD) threshold of r2 < 0.1. This was done using the LD clumping function in Plink v1.9 ^23^, with LD statistics obtained from the 1000 Genome Project. Independent significant SNPs were considered novel if they were not present in the original disorder GWAS.

We used MAGMA within the FUMA framework (v1.5.6) ^24^ to map SNPs to genes based on their position (within 35kb upstream and 10kb downstream of each gene). We performed genome-wide gene-based association tests using MAGMA. The gene-based test combines results from multiple SNPs within a gene to assess the association between the gene and the disorder, while accounting for LD between SNPs. LD information was obtained from the 1000 Genomes Phase 3 EUR reference panel, and Bonferroni correction was applied to identify genes with genome-wide significance.

We used MAGMA to conduct tissue-specific gene-set analysis and gene property analysis. Gene-set analyses assessed whether genes within an annotated set exhibited stronger associations with the disorder compared to other genes. Meanwhile, the tissue specificity test examined the relationship between tissue-specific gene expression profiles and disorder-gene associations. The gene-set analyses were performed using curated gene sets and Gene Ontology (GO) terms obtained from the Molecular Signatures Database v2023.1Hs. For the MAGMA gene property analysis, tissue expression profiles were obtained from GTEx v8 (comprising 54 tissue types) and BrainSpan (brain samples at 11 general developmental stages), available in FUMA. Gene sets and tissues were considered significant if the p-value was <0.05 after Bonferroni correction.

### Genetic correlations

We used LDSC ^4^ to compute genetic correlations between the 11 psychiatric disorders after accounting for transdiagostic effects, as well as between each of the 11 psychiatric disorders –uncorrected and corrected for p– and 34 external traits. The LD scores used were computed using 1,215,002 SNPs present in the HapMap 3 reference panel, excluding the MHC region on chromosome 6. The differences between genetic correlations with external traits before and after partialling out genetic effects associated with the p factor were assessed using a two-stage method described in Coleman et al. (2020) ^25^. In this method, first, differences in genetic correlations were assessed using a two sample z-test, and significant differences (p < 0.05) were then compared using a block-jackknife correction. The results using the jackknife were then further corrected using the Benjamini-Hochberg False Discovery Rate (FDR) method to account for multiple testing.

## Results

### Isolating transdiagnostic genetic signal from 11 major psychiatric disorders

We used Genomic SEM to construct a genomic p factor using the most recent publicly available summary statistics from GWAS of 11 major psychiatric disorders (see Supplementary Table 1). We applied a standard set of quality control (QC) filters to the 11 GWAS summary statistics within Genomic SEM and then used them in a multivariable version of LDSC. Genetic correlations are presented in Supplementary Table 2. We found a positive manifold of genetic correlation (r_G_) among most disorders, with a mean r_G_ of 0.29. Estimates ranged between -0.11 for the r_G_ between obsessive compulsive disorder (OCD) and attention deficit hyperactivity disorder (ADHD) and 0.90 between anxiety and major depressive disorder (MDD).

To capture these transdiagnostic genetic effects across all 11 disorders, we fitted a common factor model to the genetic covariance matrix. In this model, all disorders loaded on a single common factor (i.e., the p-factor; top half of Fig. 1A). This same model also allowed us to capture residual genetic variance that was associated with each disorder independent of transdiagnostic effects (i.e., non-p; bottom half of Fig. 1A). We simultaneously ran GWAS on p and residual variance in each psychiatric disorder. Figure 1A provides a diagram of the GWAS that we ran to capture genetic variance in PTSD after accounting for transdiagnostic genetic effects. We repeated this procedure 11 times to isolate transdiagnostic genetic effects from each of the 11 major psychiatric disorders. After accounting for p, we identified genome-wide significant lead SNPs for schizophrenia (SCZ, 118 hits), bipolar disorder (BIP, 22 hits), major depressive disorder (MDD, 14 hits), attention-deficit /hyperactivity disorder (ADHD, 12 hits), anorexia nervosa (AN, 10 hits), problematic alcohol use (ALCH, 3 hits), and Autism spectrum disorder (ASD, 2 hits); see Supplementary Figures 1-11, see Supplementary Tables 3-9 and Supplementary Note 4 for novel SNPs.

We used MAGMA ^26^ (see Methods) to evaluate the genetic effects of the 11 non-p GWASs on protein-coding genes. We performed gene-set analyses to identify biological pathways linked to genes associated with each major psychiatric disorder before and after accounting for p, and to analyse tissue type enrichment (Supplementary Note 4). The full results are reported in the Supplementary Notes 4 and Supplementary Tables 10-47.

### SNP heritabilities for disorders independent of p

We used LDSC implemented in Genomic SEM to estimate SNP-based heritability (h^2^) for the 11 disorders before and after partialing out the genetic variance associated with p. Figures 1B and 1C, and Supplementary Table 48 show the SNP-based h^2^ estimates for the 11 disorders (on the liability scale; Figure 1B) and for the residual variance in each psychiatric disorder after accounting for the genetic effects associated with p (on the observed scale; Figure 1C). In general, the pattern of SNP h^2^ remained similar before and after controlling for p. For example, the highest SNP h^2^ were observed for OCD and TS (.49 and .49, respectively) and the lowest estimates for ALCH and PTSD (.02 and .04).

### Removing transdiagnostic genetic effects significantly changed genetic relationships between psychiatric disorders

Figure 2 compares genetic correlations between each disorder and the other 10 disorders before and after removing the genetic variance each has in common with p (see Supplementary Figure S2 for the full genetic correlation matrices as heat maps and Supplementary Table 2 and 4 for the genetic correlation estimates and confidence intervals.) It can be seen from Figure 2 that genetic correlations were lower after accounting for genomic p. The average genetic correlation dropped from 0.29 to -0.05 after residualizing genomic p. This reduction in genetic correlation suggests greater specificity in that the pervasive contribution of p to the positive manifold of genetic correlations is reduced.

**Figure 2.**
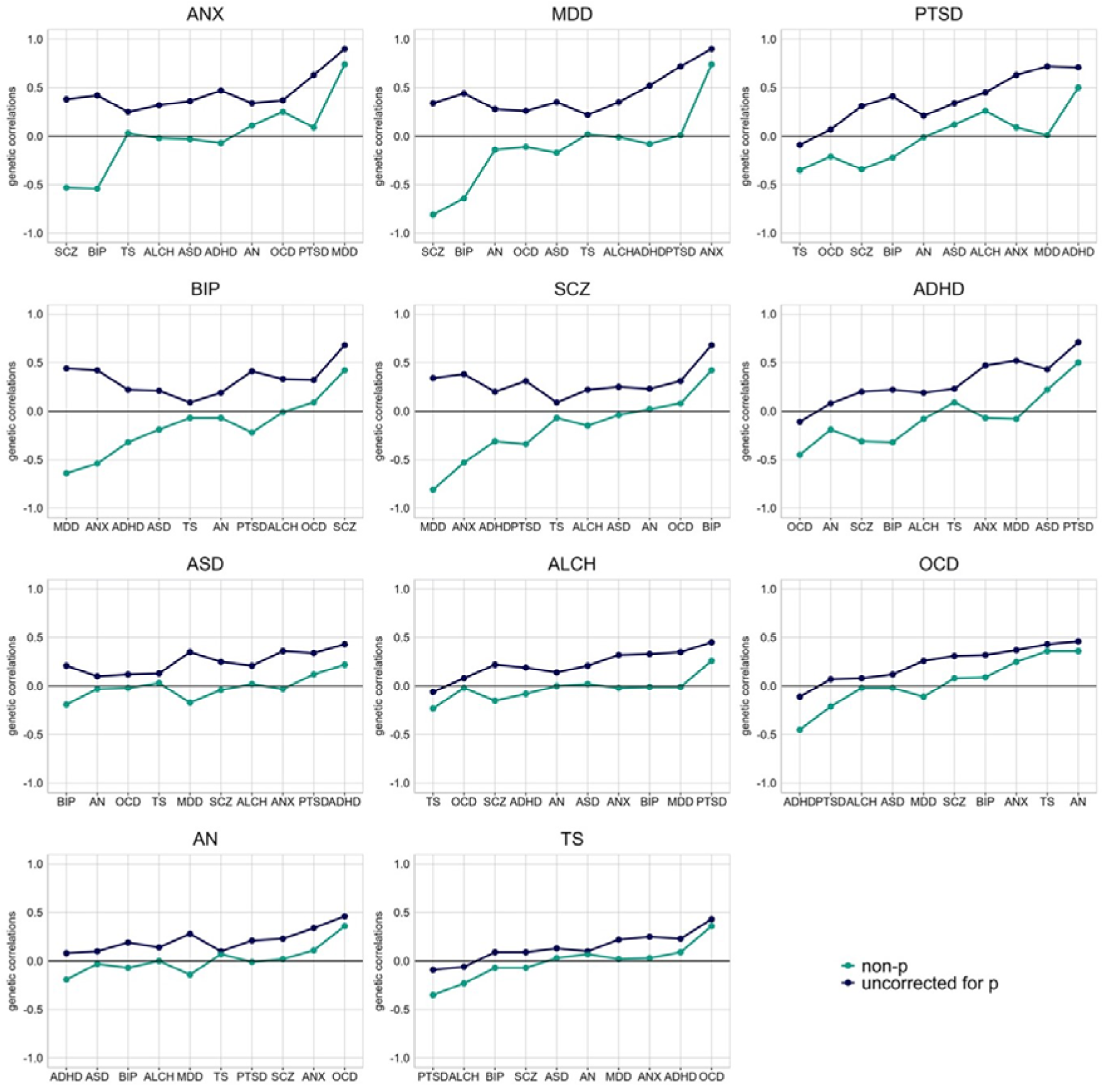
Changes in the landscape of genetic correlaitons between psychiatric disorders after accounting for transdiagnostic effects. For every psychiatric disorder, we present genetic correlations with the other 10 psychiatric conditions uncorrected for p (blue line) and the genetic correlations wit the other psychiatric disorders after removing the genetic variance captured by the p factor (non-p; green line). Disorders are presented ordered by their loading on the p factor, starting from the highest-loading disorders in the top row (i.e., PTSD, MDD and ANX) to those that shared the least amount of genetic variance with the p factor in the bottom row (i.e., AN and OCD). Correlations were estimated using LDSC within Genomic SEM. ANX, anxiety disorder; MDD, major depressive disorder; PTSD, post-traumatic stress disorder; BIP, bipolar disorder; SCZ, schizophrenia; ADHD = attention-deficit hyperactivity disorder; ASD, autism spectrum disorder; ALCH, problematic alcohol use; ; OCD, obsessive-compulsive disorder; AN, anorexia nervosa; TS, Tourette syndrome.

The major finding visualized in Figure 2 is that the reductions in transdiagnostic correlations are not uniform – that is, the blue and green lines are not parallel. For example, consider the genetic correlations between ADHD corrected and uncorrected for p with PTSD and MDD. Uncorrected for p, the genetic correlation between ADHD and PTSD is very high, 0.72. When ADHD and PTSD are corrected for p, the genetic correlation declines to 0.50, a nonsignificant difference. This means that the substantial genetic correlation between ADHD and PTSD is only slightly due to p, which could motivate the search for common mechanisms and transdiagnostic interventions. In contrast, for MDD, the genetic correlation with ADHD drops from 0.52 to -0.08. In other words, the substantial genetic correlation between ADHD and MDD disappears when ADHD is corrected for p, suggesting that the ostensible genetic relationship between ADHD and MDD is completely mediated by p.

These differences in genetic correlations are not simply due to the disorders’ loading on the genomic p factor. For example, loadings on the genomic p factor are similar for PTSD (0.78) and MDD (0.82), but the p-corrected genetic correlation with ADHD is much reduced for MDD (from 0.52 to -0.08) but not for PTSD (0.72 to 0.50). These predictive profile differences reveal differences in disorder-specific genetic architecture when the positive manifold of transdiagnostic effects of genomic p is removed.

The most interesting illustrations of prediction profile differences are cases where the genetic correlations between pairs of disorders goes from positive to negative. For example, the genetic correlation between BIP and MDD uncorrected for p was substantially positive (0.44), but after controlling for p, the genetic correlation became negative (-0.64).

Like ADHD and PTSD, for some pairs of disorders, genetic correlations remained substantial after accounting for transdiagnostic effects. For example, moderate to strong genetic correlations between ANX and MDD (rG = 0.74, SE = 0.08) and between SCZ and BIP (rG = 0.42, SE = 0.04) could still be observed. Like ADHD and MDD, for other pairs of disorders, genetic associations were not significant after accounting for transdiagnostic genetic effects. This could be observed most notably for the otherwise strong correlation between PTSD and ANX (rG = 0.63, SE = 0.08), which was reduced to 0.09 (SE = 0.17) after removing the genetic variance they shared with p. Similarly, the moderate genetic correlation between ASD and ANX (rG = 0.36, SE = 0.05) dropped to -0.03 (SE = 0.10).

Another pattern of change was observed for the genetic correlations between OCD and ADHD and between TS and PTSD. For these disorder pairs, associations that were previously negative, but small or not significant, remained negative, but their effect size increased substantially. The genetic correlation between OCD and ADHD increased from -0.11 (SE = 0.06) to -0.45 (SE = 0.09), and the genetic correlation between TS and PTSD increased from -0.09 (SE = 0.10) to -0.35 (SE = 0.19). The most dramatic pattern of change emerged for the psychotic disorders of MDD, BIP and SCZ, in which genetic correlations switched from positive to negative after removing transdiagnostic genetic effects. For MDD and SCZ, the genetic correlation changed from 0.34 (SE = .03) to -0.81 (SE = 0.05). The change was similarly dramatic for the association between MDD and BIP, from 0.44 (SE = 0.03) to -0.64 (SE = 0.06).

The same pattern of genetic correlations was obtained when isolated transdiagnostic effects and obtained disorder-specific summary statistics using a different method that followed a two-step procedure: 1) we created a genomic p factor and 2) we used GWAS-by-subtraction ^27^ to separate genetic effects associated with the genomic p-factor constructed at Step 1 from the genetic effects associated with each psychiatric disorder. (See Supplementary Note 5).

### Genetic architecture of the associations with external traits

In addition to removing transdiagnostic genetic effects from the relationships between psychiatric disorders, we also compared genetic correlations between each of the 11 psychiatric disorders – uncorrected and corrected for p – and 34 traits that are not psychiatric disorders. We focused on four broad categories of external traits: socio-demographic, anthropometric, health-related, and psychological traits. These correlations are shown in Figure 3 and 4 and Supplementary Figures 26 and 27, with details in Supplementary Table 50.

**Figure 3.**
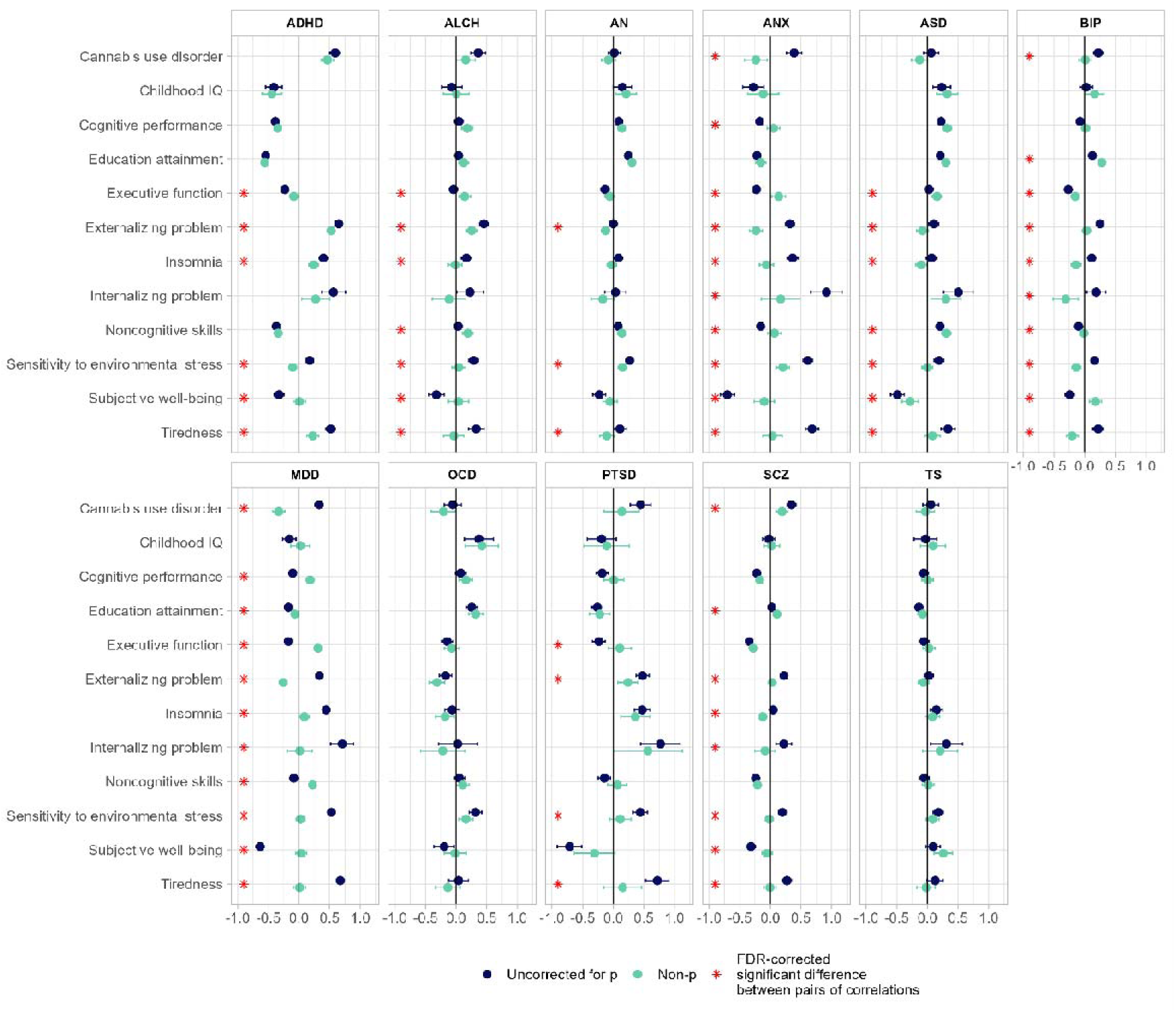
Genetic correlations between 11 major psychiatric disorders and psychological traits. The dots represent genetic correlations estimated using LDSC regression. Correlations with psychiatric disorders uncorrected for p (‘original’) are in blue, with psychiatric disorders corrected for p (‘non-p’) in green. Error bars represent 95% confidence intervals. Red asterisks indicate a statistically significant (FDR-corrected *P* < 0.05, two-tailed test) differences in the magnitude of the correlation with disorders uncorrected for p versus disorders corrected for p. Exact *P* values for all associations are reported in Supplementary Table 50. The FDR correction was applied based on all genetic correlations tested (including those reported in Supplementary Figures 26-27). Source GWASs are listed in Supplementary Table 51.

**Figure 4.**
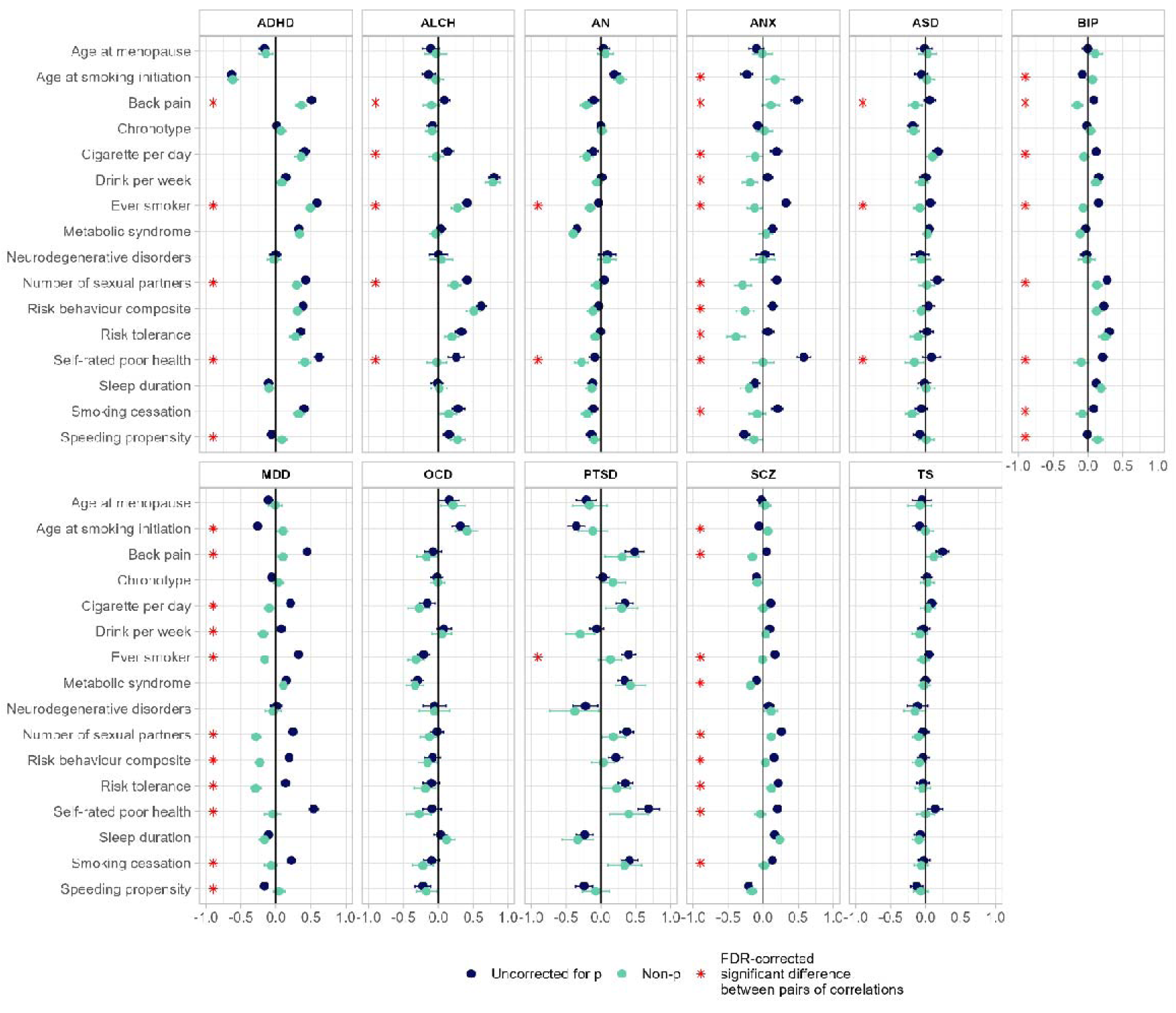
Genetic correlations between 11 major psychiatric disorders and health-related traits. The dots represent genetic correlations estimated using LDSC regression. Correlations with psychiatric disorders uncorrected for p (‘original’) are in blue, with psychiatric disorders corrected for p (‘non-p’) in green. Error bars represent 95% confidence intervals. Red asterisks indicate a statistically significant (FDR-corrected *P* < 0.05, two-tailed test) differences in the magnitude of the correlation with disorders uncorrected for p versus disorders corrected for p. Exact *P* values for all associations are reported in Supplementary Table 50. The FDR correction was applied based on all genetic correlations tested (including those reported in Supplementary Figures 26-27).

Figure 3 presents genetic correlations for the 11 psychiatric disorders uncorrected and corrected for p and psychological traits. Correlations between disorders uncorrected and corrected for p and most psychological traits differed significantly, as indicated by the red asterisks. In almost all cases, the genetic correlations were lower for the corrected than for the uncorrected psychiatric traits. Importantly, correlations with the same traits often appeared to be significantly different across major psychiatric disorders, such as subjective wellbeing, sensitivity to environmental stress and tiredness. For example, consider sensitivity to environmental stress. Uncorrected for p, genetic correlations were significant and substantial between sensitivity to environmental stress and major psychiatric disorders, but corrected for p, these correlations were significantly lower and sometimes negligible: SCZ (0.19 vs -0.02), MDD (0.53 vs 0.03), ASD (0.19 vs. -0.002) and ALCH (0.29 vs. 0.04). A similar pattern of results was observed for several external traits that often co-occur with most psychiatric disorders, such as subjective wellbeing, loneliness, tiredness and insomnia.

In other words, genetic effects associated with each major psychiatric disorder also include overlapping genetic variance associated with these transdiagnostic traits, such as sensitivity to environmental stress, which mask their specificity. Supplementary Figures 26-27 and Supplementary Table 50 present the genetic correlations between psychiatric disorders and all socio-demographic, anthropometric, health-related and psychological traits.

As shown in Figure 4, similar results emerged for health-related traits. Genetic correlations between disorders corrected for p were generally lower than those uncorrected for p, often significantly lower. For example, self-reported poor health showed significant reductions for most disorders (ADHD, ALCH, ANX, BIP, MDD, SCZ); number of sexual partners showed significant reductions for four disorders (ADHD, ALCH, BIP, SCZ); and cigarette per day yielded significant reductions for three disorders (ALCH, ANX, MDD; see Supplementary Table 50).

Though most of the changes in correlations were reductions, there were some exceptions. For example, after accounting for p, the genetic correlation between BIP and SCZ and educational attainment increased significantly. Other genetic correlations reversed after accounting for p. For example, the genetic correlation between BIP and loneliness changed from r_G_ = 0.11 before accounting for p to r_G_ = -0.26 after removing transdiagnostic effects, a similar change was also observed for the genetic correlations between BIP and Back Pain (from 0.08 to -0.15) and MDD and several cognitive traits (cognitive performance (from -0.10 to 0.18), executive functions (from -0.18 to 0.31) and noncognitive skills (from -0.09 to 0.22) and risk-taking behaviours (e.g., the correlation between MDD and risk tolerance changed from 0.14 to -0.29). Significant changes were also observed for the genetic correlations between ANX and risk-taking behaviour (from 0.14 to -0.26), ANX and risk tolerance (from 0.07 to -0.39), and SCZ and back pain (from 0.05 to -0.16). Removing transdiagnostic genetic effects from psychiatric disorders, particularly MDD, ANX, SCZ and BIP, resulted in divergent patterns of genetic associations with external biobehavioural traits.

## DISCUSSION

One of the most important findings from the genomic analysis of psychiatric disorders is the consistent evidence for genetic overlap among disorders ^10–12^, leading to the concept of a general factor called genomic p. While acknowledging the importance of p, the present paper’s focus on genetic specificity is motivated by the need to control for the pervasive transdiagnostic influence of p in order to understand the genetic architecture of psychiatric disorders independent of p. The results show that the genetic landscape shifts dramatically when we remove the effect of genomic p from each of 11 psychiatric disorders.

A striking example is the genetic relationship between Bipolar Disorder and Major Depressive Disorder. As has been found in other studies, uncorrected for p, the genetic correlation is substantially positive, 0.44 in our analysis. However, after controlling for p, the genetic correlation was highly negative (-0.64). This negative correlation suggests that, once transdiagnostic effects are accounted for, a genetic liability for one disorder conveys a lower genetic liability for the other disorder. This finding has the potential to inform diagnostic categorization and has far-reaching implications for future studies aimed at understanding biological causes and treatments for these disorders. The positive genetic correlation between BIP and MDD uncorrected for p would suggest a search for common mechanisms and transdiagnostic interventions that affect BIP and MDD similarly. However, when p is controlled, their strong *negative* genetic correlation suggests that mechanisms and interventions work in opposite directions for BIP and MDD.

The pattern of genetic correlations between several other disorders also shifted dramatically for other psychiatric disorders, highlighting discrepancies between diagnostic nosology and genetic structure of psychiatric disorders. For some pairs of disorders, positive genetic correlations disappeared entirely after correction for p, such as the genetic correlation between ANX and PTSD, suggesting that their overlap is entirely captured by what cuts across all psychiatric diagnoses. Another major shift was observed for the slightly negative genetic correlation between OCD and ADHD, which became strongly negative after accounting for p. This suggests that their genetic overlap with p obscured the negative genetic relationship between these disorders. This finding is consistent with the current clinical perspective suggesting that OCD and ADHD lie at the opposite extremes of the impulsivity-compulsivity continuum ^28, 29^.

Other pairs of disorders remained correlated positively after accounting for p, such as the genetic correlation between SCZ and BIP and between ANX and MDD, which suggests that only part of their overlap is shared with the other disorders included in our transdiagnostic model. It is important to remember that, although latent variables serve to summarize patterns of comorbidity or covariation among indicators, they are statistical constructs that depend on the indicators that are included in each model.

Removing transdiagnostic genetic effects from psychiatric disorders, particularly MDD, ANX, SCZ and BIP, also resulted in divergent patterns of genetic associations with external biobehavioural traits, especially health-related and psychological traits. In other words, genetic effects associated with each major psychiatric disorder also include overlapping genetic variance associated with these transdiagnostic traits, such as sensitivity to environmental stress, which mask their specificity. Focusing on disorder-specific genetic effects provides novel insights into the genetic architecture, biology and comorbidity between psychiatric conditions that can inform research on sequelae and antecedents as well as treatment.

Together, our findings highlight how isolating transdiagnostic genetic risk from major psychiatric disorders provides novel insight into disorder-specific genetic architecture and a more nuanced understanding of their comorbidities and co-occurrences with psychological and health-related traits. Consequently, these findings emphasize the significance of considering specificity as well as generality in psychiatric genetics. By demonstrating distinct genetic correlations and outcomes associated with psychiatric conditions independent of transdiagnostic effects, the findings pave the way for new avenues of research. One such application is the use of non-p summary statistics to create polygenic scores that index greater specificity in psychiatric disorders (Keser et al., in press). For example, isolating transdiagnostic effects from psychiatric polygenic risk scores can lead to refined predictions of how biological risk for each disorder unfolds developmentally and can offer new avenues to investigate how genetic and environmental risk combine.

Our findings need to be interpreted in the context of their limitations. The most important limitation is that our research begins with case-control GWAS based on traditional diagnoses perfused with transdiagnostic effects. This points to the need for GWA research using phenotypes that correspond more closely to the genetic architecture of psychopathology ^1^. The origin of psychiatric nosology is historical rather than empirical; progress depends on more empirically derived dimensional approaches such as HiTop ^30^ and Rdoc ^31^.

Another limitation is that p is a statistical construct for which there is no consensus on what it is or how to measure it ^32^, which leaves non-p even further adrift from reality. However, similar accusations could be levelled at g, the general factor that emerges from diverse cognitive traits, but g is one of the most stable and predictive variables in the behavioural sciences ^33^. g is what diverse cognitive traits have in common and is not caused by any single physiological process, such as speed of neural conduction, nor is it defined by any single psychological process such as abstract reasoning. We embrace the possibility that p, like g, is not one thing – it is precisely what diverse traits have in common ^34^. We suggest that p will be similarly valuable for understanding general genetic influences, and that isolating the transdiagnostic effects captured by p will be useful in sharpening research on specific genetic influences, particularly in the context of developmental psychopathology and clinical epidemiological studies. Genetic effects that are disorder-specific might inform future research into causes and consequences of psychiatric conditions applying causal designs including mendelian randomization and longitudinal models ^35, 36^.

A further limitation is related to our choice of modelling p as a common factor, given our interest in capturing transdiagnostic genetic effects that could index shared genetic liability across all 11 major psychiatric disorders. Although alternative models have been proposed ^10, 17, 37^ different statistical approaches to modelling p were found to lead to similar estimates ^17^. Relatedly, we allowed our indicators to load freely onto the common factor, as such, some disorders (e.g., ANX, MDD and PTSD) contributed more than others to the general factor (e.g., OCD and AN). A model in which all indicators are restricted to contribute the same amount of variance to the general factor would likely have led to different results, although arguably it would have provided a poorer account of transdiagnostic effects in psychopathology. It should also be noted that, while general factor models can fit psychopathological data, alternative explanations have been proposed, most notably network models ^38^.

Other limitations are general issues in GWA research. For example, the GWA studies that are the basis for this research are largely limited to individuals of European ancestry, so the results reported here might not generalize beyond this population ^39, 40^. In addition, the contributing GWA studies are meta-analyses of different cohorts that may be subject to heterogeneity that cannot be fully quantified. Moreover, our research is limited by issues that could affect any GWA results, such as cross-trait assortative mating ^41^ and population stratification ^42^.

In conclusion, our results show that isolating transdiagnostic effects from major psychiatric disorders provides novel insight into disorder-specific genetic architecture by providing. a more precise understanding of comorbidities and co-occurrences in psychopathology. Until better correspondence between psychiatric diagnoses and the genetic architecture of psychopathology is achieved, isolating p from diagnostic categories will sharpen genetic research by focusing on disorder-specific genetic effects.

## Data Availability

The data is publicly available online. All data produced will be made available online upon publication at the following link: https://github.com/CoDEresearchlab

## Acknowledgements

E.K. is supported by a PhD scholarship from the Economic and Social Research Council. W.L. is supported by a Chinese Scholarship Council PhD fellowship. MM is supported by a starting grant from the School of Biological and Behavioural Sciences at Queen Mary University of London. We would like to thank Jonathan Coleman for sharing the scripts used to apply block-jackknife correction, which were written by Héléna Gaspar and Christopher Hübel.

## Contributions

E.K., W.L., R.P. and M.M. conceived and designed the study.; E.K. and W.L. analyzed the data with contributions from M.M. and A.G.A.. E.K., W.L., R.P. and M.M. wrote the paper with contributions from K.R., A.G.A., and T.C.E. All authors contributed to the interpretation of data, provided critical feedback on manuscript drafts, and approved the final draft.

## Code availability

Code will be available at https://github.com/CoDEresearchlab upon publication. Ahead of publication we will make the code available upon request.

