## Supplementary Notes and Figures for "Isolating transdiagnostic effects reveals specific genetic profiles in psychiatric disorders"

1. Extensions of preregistered analyses ………………………………………………………………………………..3

2. Genomic Structural Equation Modelling ……………………………………………………………………………3

Processing of summary statistics ……………………………………………………………………………..3

#### 3. A common factor model to capture transdiagnostic effects ………………………………..............4

Effective sample size calculation ................................................................................ 4

4. Novel genetic variants associated with psychiatric disorders beyond transdiagnostic effects and MAGMA results………………………………………………………………………………………………….5

#### 5. An alternative approach to identify genetic variants associated with psychiatric disorders after accounting for transdiagnostic effects using GWAS-by-subtraction …………………………….6

Supplementary Figure 19. The brain sample enrichment results on 11 general brain developmental stages by BrainSpan of ADHD uncorrected and corrected for p………………….28

Supplementary Figure 290. The tissue type enrichment results on 53 specific tissue types by GTEx of ASD uncorrected and corrected for p .......................................................................29

Supplementary Figure 21. The brain sample enrichment results on 11 general brain developmental stages by BrainSpan of ASD uncorrected and corrected for p…………………….30

Supplementary Figure 22. The tissue type enrichment results on 53 specific tissue types by GTEx of ALCH uncorrected and corrected for p .....................................................................31

Supplementary Figure 23. The brain sample enrichment results on 11 general brain developmental stages by BrainSpan of ALCH uncorrected and corrected for p…………………..32

Supplementary Figure 24. The tissue type enrichment results on 53 specific tissue types by GTEx of AN uncorrected and corrected for p………………………………………………………………………33

Supplementary Figure 25. The brain sample enrichment results on 11 general brain developmental stages by BrainSpan of AN uncorrected and corrected for p……………………..34

Supplementary Figure 26. Genetic correlations between the psychiatric disorders and anthropometric traits before and after controlling for p ......................................................35

Supplementary Figure 27. Genetic correlations between psychiatric disorders and socio-demographic traits before and after controlling for p ..........................................................36

### SUPPLEMENTARY NOTES

#### Supplementary Note 1: Extensions of pre-registered analyses

We extended our pre-registered analytical plan by including an alternative approach to isolating transdiagnostic genetic effects from each psychiatric disorders, this approach, based on a single common factor model is now presented in the main manuscript, whilst the modelling approach proposed in our pre-registered analyses is presented in Supplementary Note 5. Results were highly consistent across both modelling approaches.

#### Supplementary Note 2: Genomic Structural Equation Modelling

Genomic structural equation modelling (Genomic SEM ^1^) is a framework that applies structural equation modelling to GWAS summary statistics to model patterns of genetic correlations between complex traits.

Genomic SEM uses a two-stage estimation process ^1^. In the first stage, the genetic covariance matrix and associated sampling covariance matrix are estimated. In the second stage, a model is specified, and parameters are estimated by attempting to minimize the discrepancy between the model-implied genetic covariance matrix and the empirical covariance matrix. The fit of the model can then be evaluated using standard goodness-of-fit indices, including standardized root mean square residual (SRMR), model *χ*2, Akaike Information Criterion (ACI) and the Comparative Fit Index (CFI).

##### Processing of summary statistics

We processed GWAS summary statistics for use in Genomic SEM as follows. Summary statistics were formatted with the *munge* function within Genomic SEM R package v.0.0.5 (using default parameters). The *munge* function converts the summary statistics to the format expected by LDSC by removing all the SNPs that are not present in the reference panel, and, when the information is available, filtering out SNPs with minor allele frequency < 1% and information score < 0.9. The HapMap3 reference file is provided in the Genomic SEM repository (<https://github.com/GenomicSEM/GenomicSEM>). The processed summary statistics were then used in a multivariable version of LDSC implemented in Genomic SEM to estimate the genetic covariance and sampling covariance matrices for the 11 disorders i.e., SCZ, BIP, ADHD, MDD, ANX, PTSD, ALCH, ASD, OCD, AN, TS. The genetic covariance matrix contains SNP-based heritability estimates on the diagonal and genetic covariance on the off-diagonal. The sampling covariance matrix contains squared standard errors on the diagonal (the sampling variances) and sampling covariances on the off-diagonal, which index sampling dependencies that will arise when there is participant sample overlap. The LD weights used for LDSC were calculated using the European subsample of the 1000 Genome Phase 3 project, excluding the major histocompatibility complex due to complex LD structures in this region that can bias estimates.

The *sumstats* function within Genomic SEM was then used to standardize all SNPs. In this step, summary statistics for each of the 11 disorders were restricted to SNPs with minor allele frequency > 1% and information score > 0.6, and to SNPs that were present for all of the 11 disorders. The summary statistics were also filtered to SNPs present in the European-only 1000 Genomes Phase 3 reference panel. After these quality control steps, 3,746,806 SNPs were present across all 11 disorders.

Before running the model, the genetic covariance and sampling covariance matrices were transformed into genetic correlation and sampling correlation matrices. These standardized LDSC matrices were then used as input into the model specified within Genomic SEM. The default diagonally weighted least squares (DWLS) estimator was used to run the model which allowed our indicators to load freely onto the common factor. We run the model using the alternative maximum likelihood (ML) estimator which restricts the indicators to contribute the same amount of variance to the general factor; however, it provided a poorer model fit hence we continued with DWLS in the model.

#### Supplementary Note 3: A common factor model to capture transdiagnostic effects

To capture transdiagnostic genetic effects across all 11 disorders, we fitted a common factor model to the genetic covariance matrix produced in Genomic SEM. In this model, all disorders loaded on a single common factor. Model fit was assessed using standard indices used in structural equation modelling, including standardized root mean square residual (SRMR), model *χ*2, Akaike Information Criterion (ACI) and the Comparative Fit Index (CFI). The model provided adequate fit, (χ^2^(44) = 950.4836, AIC = 994.48, CFI = .82, SRMR = .12). (The model fit using the ML estimator was χ^2^(44) = 1245.822, AIC = 1289.82, CFI = .77, SRMR = .13)

##### Effective sample size calculation

We calculated effective sample sizes for each of the 11 disorders following the method described by Grotzinger et al. (2023) ^2^. According to this method, the effective sample size for GWAS that was meta-analysed was calculated by summing the effective sample sizes across contributing cohorts. This approach incorporates cohort-specific ascertainment, which provided an unbiased estimate of liability scale heritability for binary traits. We refer the reader to Grotzinger et al. (2023) ^2^ for a detailed explanation of why this method produces a more accurate estimate of heritability for binary traits.

**Supplementary Note 4: Novel genetic variants associated with psychiatric disorders beyond transdiagnostic effects and MAGMA results**
After accounting for the transdiagnostic genetic effects associated with p, we identified independent significant hits for seven out of eleven GWASs (see Method). The largest number of independent hits was observed for the ‘non-p’ GWAS of schizophrenia (118, 27 of which were novel SNP associations that had not emerged as significant in the original GWAS), followed by BIP (22, 13 novel SNP associations), MDD (14, 11 novel SNP associations), ADHD (12, 8 novel SNP associations), AN (10, 8 novel SNP associations), ALCH (3, 1 novel SNP associations) and ASD (2). Some of these novel SNP associations had been uncovered by GWAS of psychiatric disorders other than the 11 included in our model, or by GWAS of other traits (not psychiatric disorders), while others had not been reported in the previous literature. For example, focusing on schizophrenia, 7 out of the 27 novel SNP associations had been uncovered by previous genomic studies on schizophrenia ^3-6^, 6 had been reported as SNPs associated with physiological or psychological traits (i.e.; body mass index (BMI) ^7, 8^ and intelligence ^9^), and 14 SNP associations had not been reported in the extant literature. Details regarding the novel SNP uncovered for the other six ‘non-p’ GWASs are presented in the Supplementary Tables 3-9.

We also used MAGMA ^10^ in FUMA to evaluate the genetic effects of the 11 non-p GWASs (see Methods). The gene-level and gene-set analyses were performed to identify biological pathways linked to genes associated with each major psychiatric disorder before and after accounting for p, and to analyse tissue type enrichment. The method is briefly described in the main manuscript and additional details can be found in Watanabe et al. (2017)^11^ and Watanabe et al (2019) ^11^. All results reported are Bonferroni's corrected to reduce multiple comparison problems. We found that after accounting for transdiagnostic genetic effects, 316 genes were associated with SCZ, 63 genes were associated with BIP, 44 with MDD, 29 with ADHD, 37 with AN, 5 with ASD, and 1 with ALCH. Only SCZ, BIP and MDD showed significant enriched gene sets after accounting for p. SCZ non-p was associated with enrichment in six gene-sets related to neuron system (Supplementary Table 11). MDD non-p was significantly associated with enrichment in 1 gene-set related to subpallium development (Supplementary Table 15). BIP non-p was significantly associated with enrichment in 1 gene-set (Supplementary Table 13). The full results are reported in the Supplementary Tables 10-19.

We tested whether common variants in genes specifically expressed in 53 Genotype-Tissue Expression (GTEx) tissues were enriched in their effects on psychiatric disorders (SCZ, BIP, MDD, ADHD, ALCH, ASD, AN) after accounting for transdiagnostic effects (See Methods). Genes predominantly expressed in the brain cortex and other brain-specific tissues were enriched in MDD, BIP, SCZ, ADHD, ASD and AN (Supplementary Figures 13-25, and Supplementary Table 12-47). Enrichment patterns were overall consistent between psychiatric disorders before and after removing transdiagnostic signals, but there were some exceptions. First, brain development stages enrichment results showed that, for SCZ, the early-to-late prenatal stages were enriched before accounting for p, but no longer enriched after removing transdiagnostic effects. Late infancy remained the most enriched developmental stage for SCZ, showing the strongest associations (Supplementary Figure 17, and Supplementary Table 21, 23). Second, for ASD, tissue property enrichment analyses showed that ten brain regions were enriched, with the substantia nigra showing the strongest signal, after accounting for the transdiagnostic effects, compared to four brain regions before (Supplementary Figure 21, and Supplementary Table 44, 46).

#### Supplementary Note 5: An alternative approach to identify genetic variants associated with psychiatric disorders after accounting for transdiagnostic effects using GWAS-by-subtraction.

##### A two-stage modelling approach

We proceeded in two stages: first, we modelled a genomic p factor using a common factor model in Genomic SEM (see Supplementary Note 2 and 3), and second, we used the GWAS-by-subtraction ^12^ approach to isolate genetic effects associated with p from those contributing to each major psychiatric disorder that were not captured by p.

We used GWAS-by-subtraction to separate genetic effects associated with the previously constructed genomic p-factor from the genetic effects associated with each psychiatric disorder. This allowed us to identify genetic effects associated with each disorder independent of transdiagnostic genetic effects. Figure S1 provides a diagram of the GWAS-by-subtraction model using SCZ as an example. We repeated this procedure 11 times to isolate transdiagnostic genetic effects from each of the 11 major psychiatric disorders.

##### GWAS-by-subtraction

GWAS-by-subtraction ^12^ is a specific model within Genomic SEM that that estimates, for each SNP, an effect on a specific trait that is independent of that SNP’s effect on another trait. Using schizophrenia as an example, the GWAS summary statistics for p factor and SCZ are regressed on a latent factor, ‘p SCZ’, which represents genetic variance that is shared across the 11 disorders (Figure S1, left). SCZ was further regressed on a second latent factor representing the residual genetic variance in SCZ left over after regressing out variance related to p, i.e., non-p SCZ (Figure S1, right). By construction, the genetic variance in non-p SCZ is independent of genetic variance in the p-factor (*r*_g_ = 0). In other words, the non-p SCZ factor represents genetic variance in SCZ that is not accounted for by the p-factor. The two latent variables, p-SCZ and non-p SCZ are then regressed on each SNP, iterating across all SNPs in the GWAS, resulting in new GWAS summary statistics for non-p SCZ.

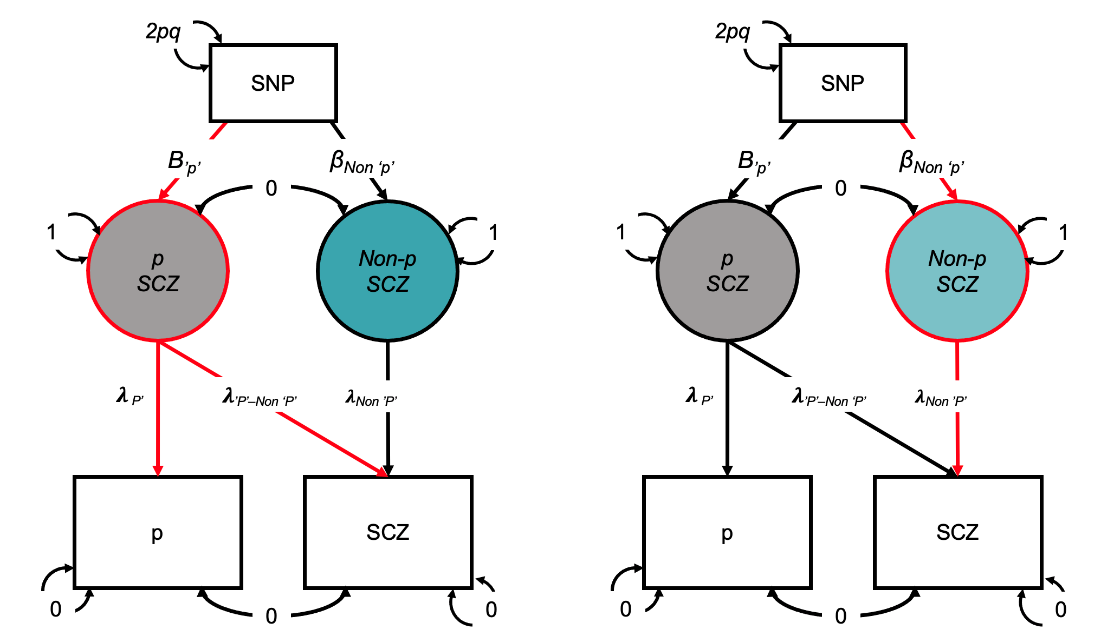

**Figure S1.** Schematic overview of the GWAS-by-subtraction approach for creating a latent residual for the schizophrenia (SCZ)-specific factor (Figure adapted from Demange et al., 2021 ^12^). In this diagram, squares represent the observed SNP and the GWAS of p-factor and SCZ. Circles represent the latent (unobserved) variables, i.e., P SCZ and Non-p SCZ. Single-headed arrows represent linear regression associations pointing from the independent variable to the dependent variable. Two-headed arrows represent covariance relationships. The covariances between p-factor and SCZ and between p SCZ and Non-p SCZ are fixed to 0. The variance of the SNP is fixed to the value of 2pq (p= reference allele frequency, q= alternative allele frequency, based on 1000 Genomes Project phase 3). The residual variances of p and SCZ are fixed to 0, so that all variance is explained by the latent factors. The variances of the latent factors are fixed to 1. λ= freely estimated factor loadings that are equivalent to regression weights. Β= regression effects of p SCZ and Non-p SCZ on the SNP.

The results obtained using this alternative approach were highly consistent with those obtained using the approach described in the main manuscript, as indicated by the changes in the genetic correlations presented in Figure S2.

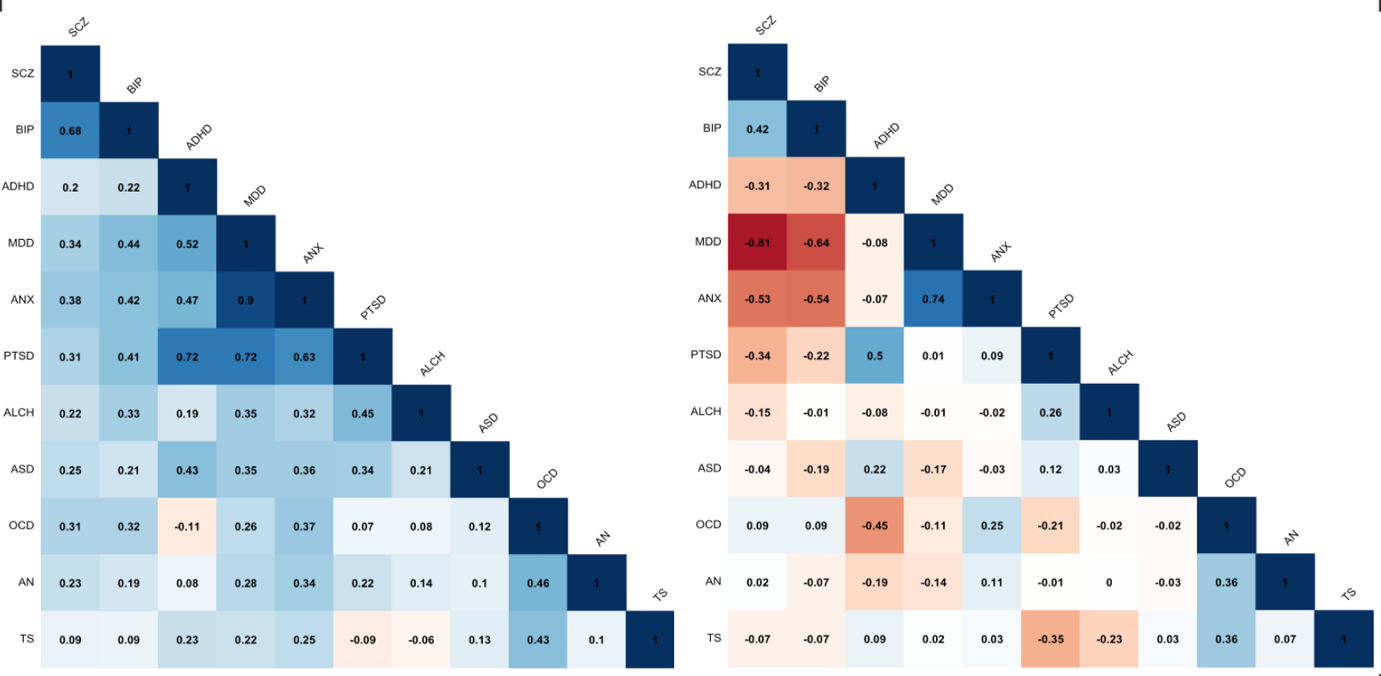

**Figure S2. Genetic correlations between psychiatric disorders before and after accounting for transdiagnostic effects obtained using the GWAS-by-subtraction modelling approach. A)** Genetic correlations between 11 major psychiatric disorders uncorrected for p. **b)** Genetic correlations between psychiatric disorders after removing the genetic variance each disorder shares with p using the two-stages approach described in Supplementary Note 5. Correlations were estimated using LDSC within Genomic SEM.

##### Effective sample size calculation for latent residual factors

We calculated the expected sample size of the latent non-p factors following the method described by Demange et al. (2021) ^12^. According to this method, effective sample sizes, estimated based on the formula described by Mallard et al. (2022) ^13^, are adjusted by multiplying them by the residual heritability (squared unstandardized path loading).

### SUPPLEMENTARY FIGURES

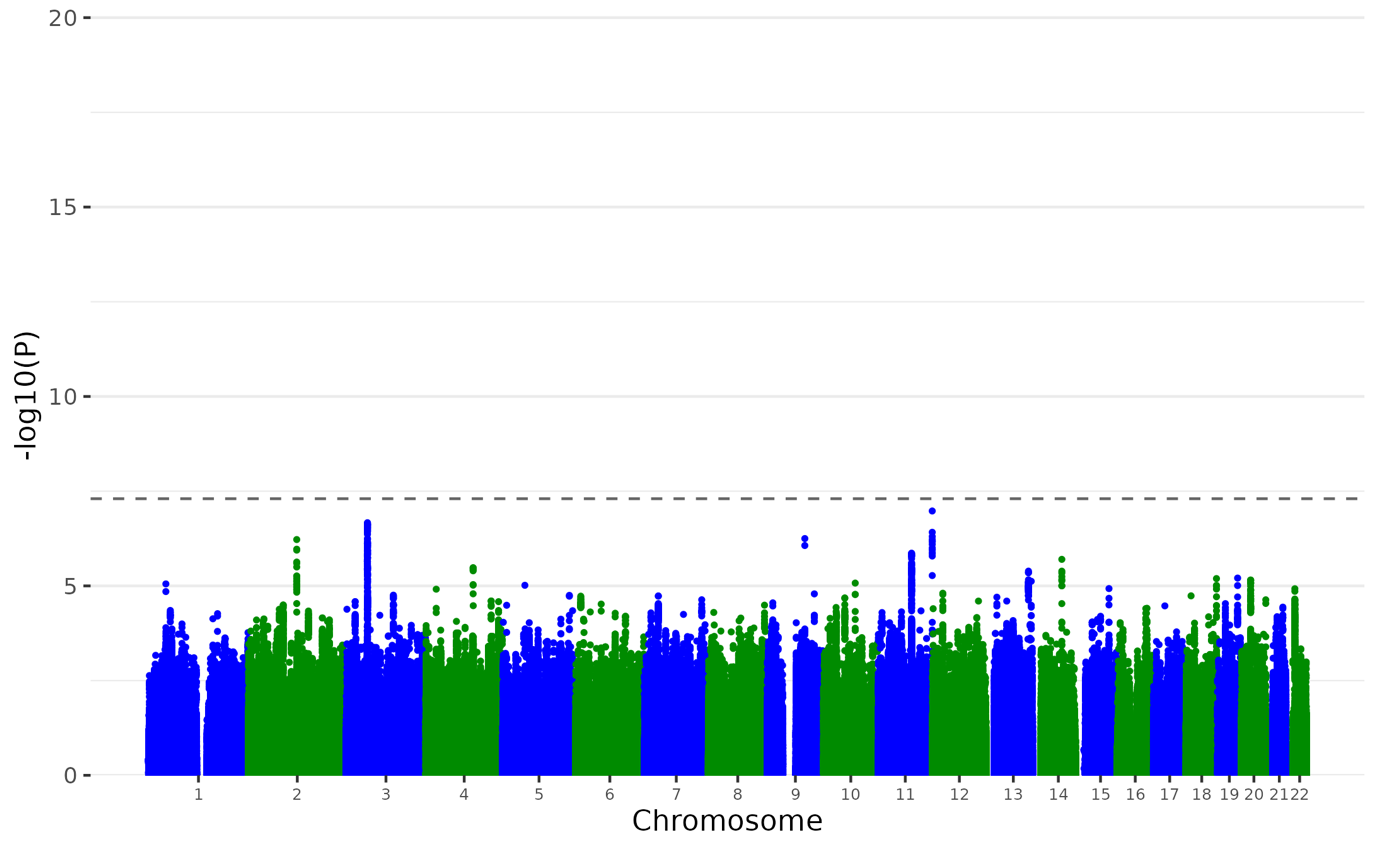

**Supplementary Figure 1. Manhattan plot of the ANX Non-p GWAS.** Plot of the -log_10_(p-value) associated with the Wald test (two-sided) of β_p_ for all SNPs ordered by chromosome and base position. Red diamonds indicate genome-wide significant independent hits (within a 250Kb window and r^2^ < .1) associations.

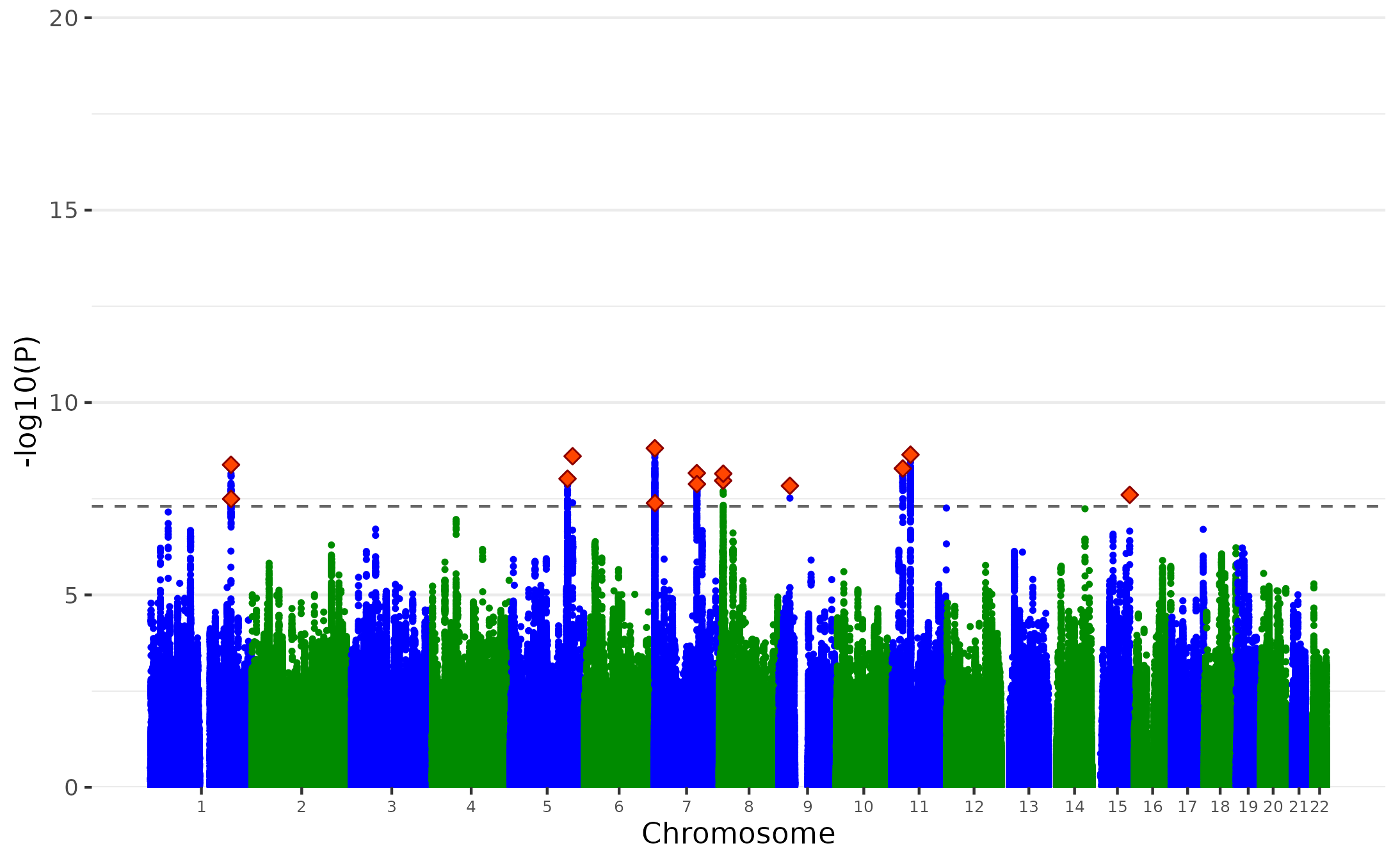

**Supplementary Figure 2. Manhattan plot of the MDD Non-p GWAS.** Plot of the -log_10_(p-value) associated with the Wald test (two-sided) of β_p_ for all SNPs ordered by chromosome and base position. Red diamonds indicate genome-wide significant independent hits (within a 250Kb window and r^2^ < .1) associations.

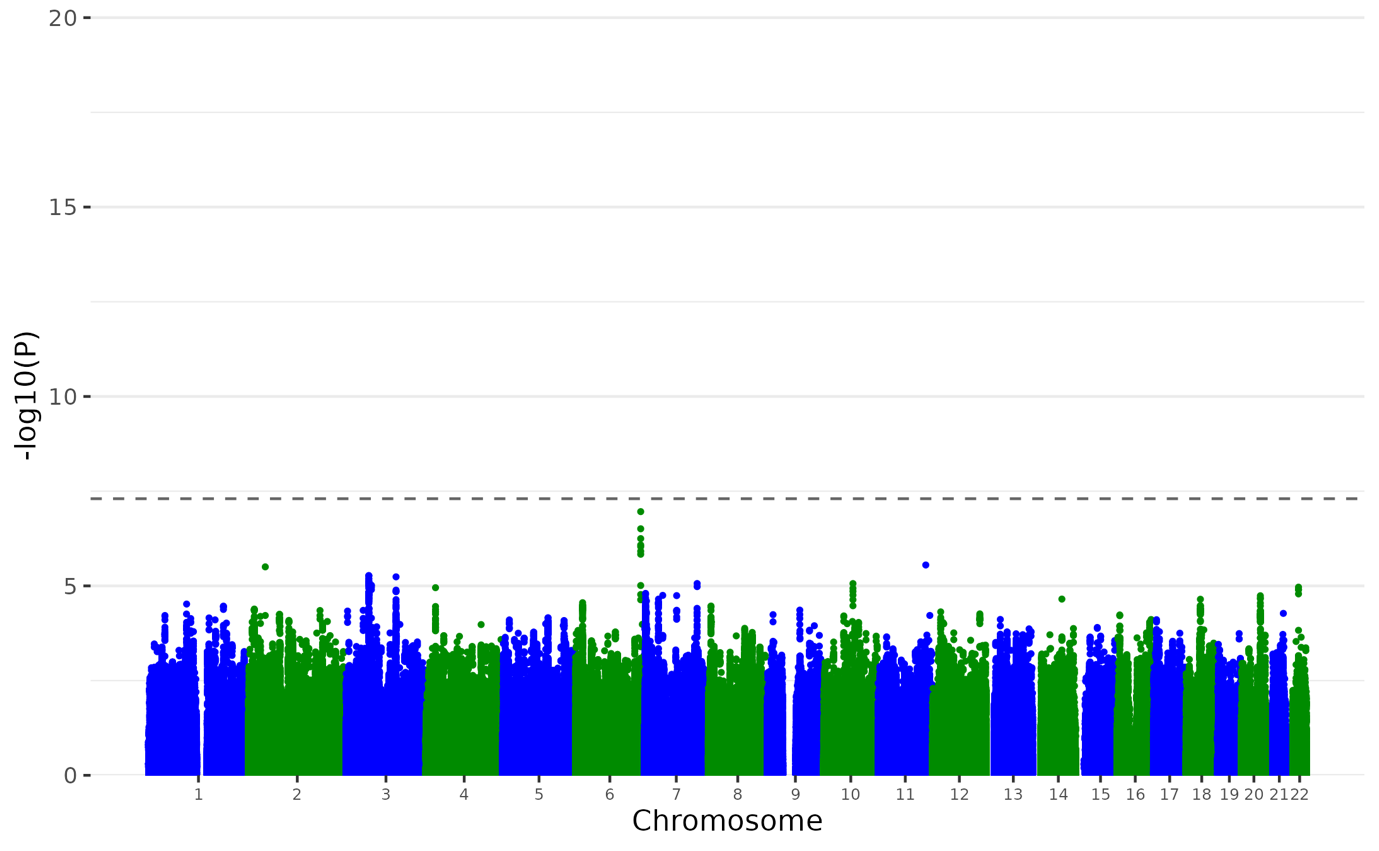

**Supplementary Figure 3. Manhattan plot of the PTSD Non-p GWAS.** Plot of the -log_10_(p-value) associated with the Wald test (two-sided) of β_p_ for all SNPs ordered by chromosome and base position. Red diamonds indicate genome-wide significant independent hits (within a 250Kb window and r^2^ < .1) associations.

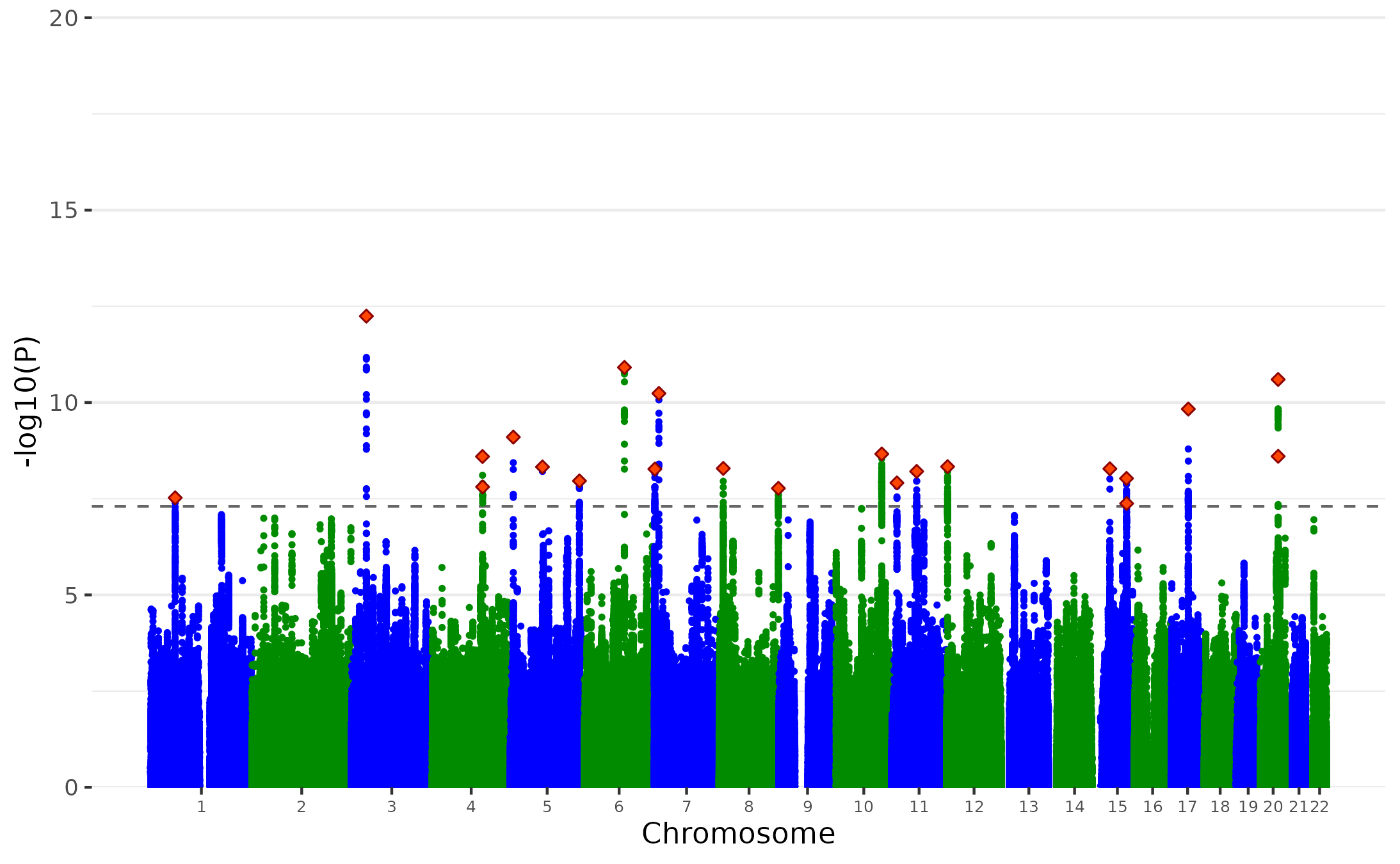

**Supplementary Figure 4. Manhattan plot of the BIP Non-p GWAS.** Plot of the -log_10_(p-value) associated with the Wald test (two-sided) of β_p_ for all SNPs ordered by chromosome and base position. Red diamonds indicate genome-wide significant independent hits (within a 250Kb window and r^2^ < .1) associations.

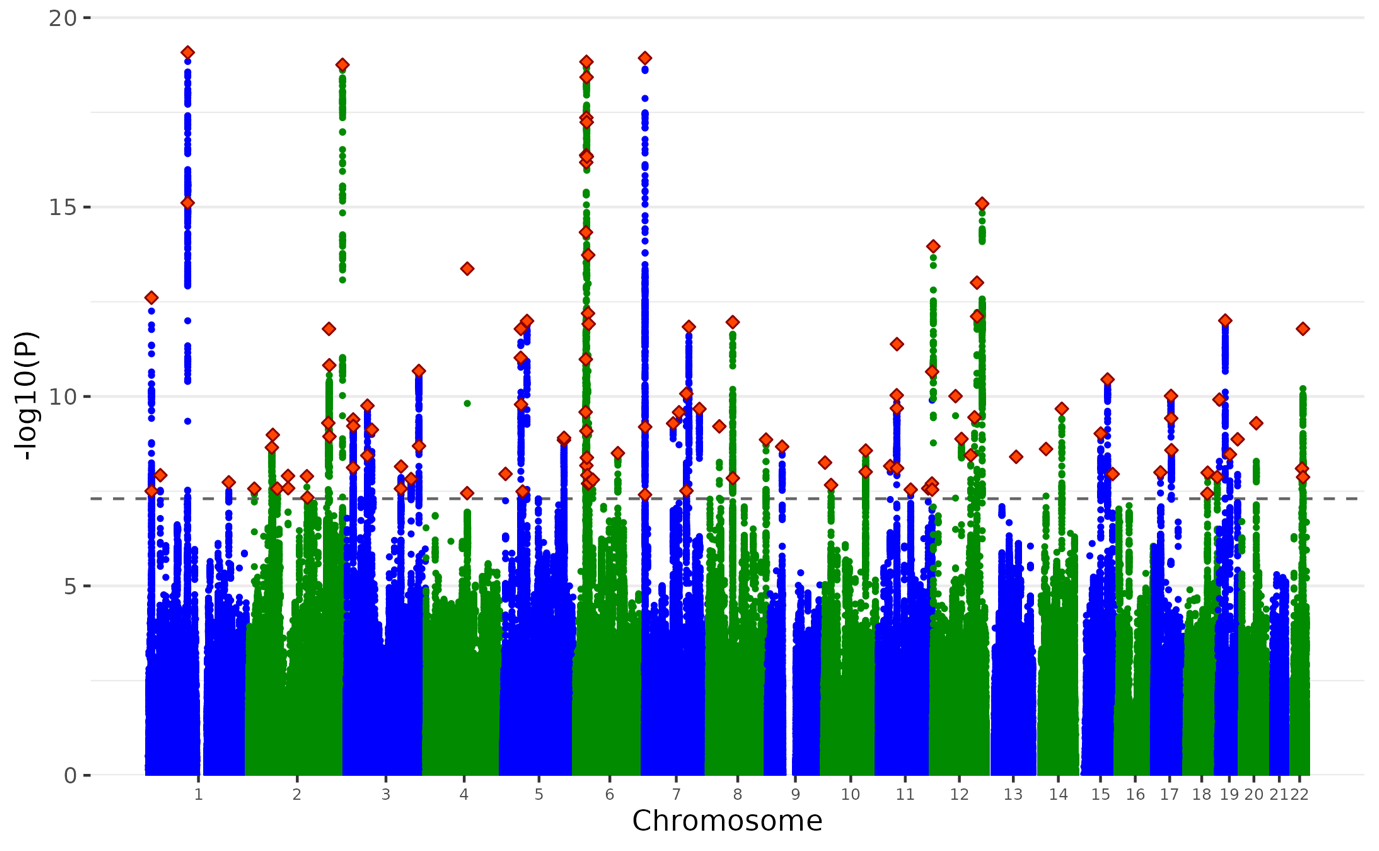

**Supplementary Figure 5. Manhattan plot of the SCZ Non-p GWAS.** Plot of the -log_10_(p-value) associated with the Wald test (two-sided) of β_p_ for all SNPs ordered by chromosome and base position. Red diamonds indicate genome-wide significant independent hits (within a 250Kb window and r^2^ < .1) associations.

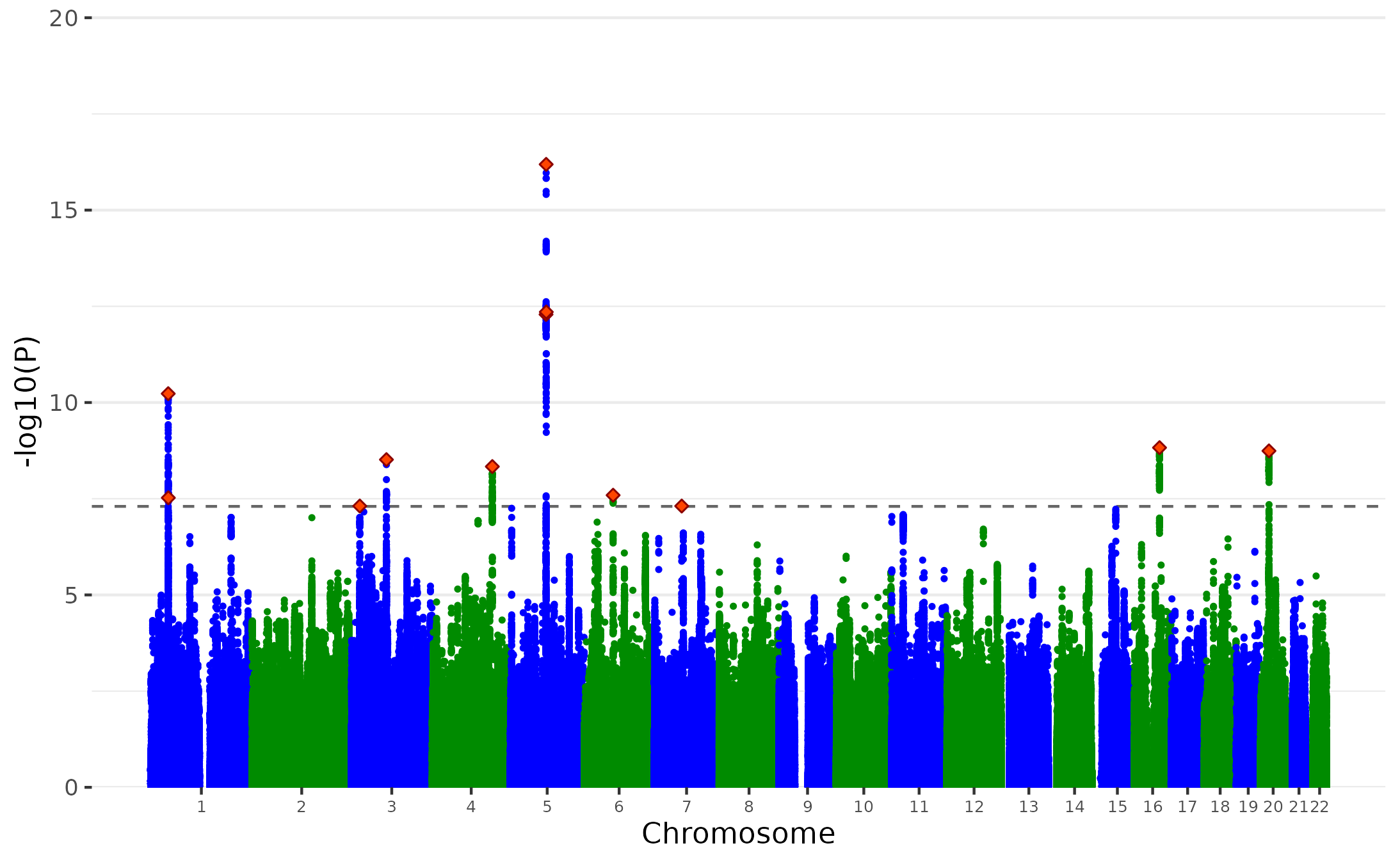

**Supplementary Figure 6. Manhattan plot of the ADHD Non-p GWAS.** Plot of the -log_10_(p-value) associated with the Wald test (two-sided) of β_p_ for all SNPs ordered by chromosome and base position. Red diamonds indicate genome-wide significant independent hits (within a 250Kb window and r^2^ < .1) associations.

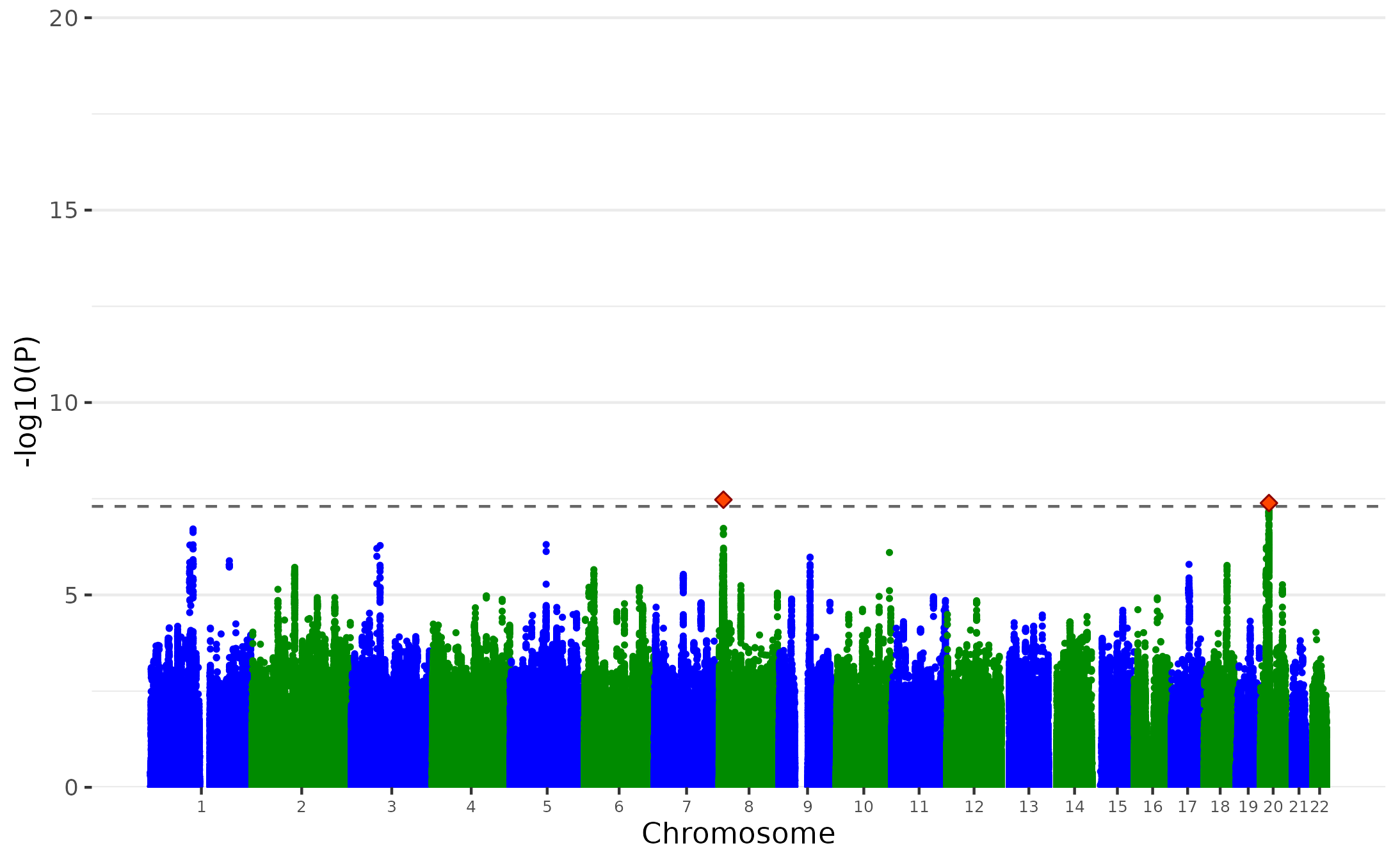

**Supplementary Figure 7. Manhattan plot of the ASD Non-p GWAS.** Plot of the -log_10_(p-value) associated with the Wald test (two-sided) of β_p_ for all SNPs ordered by chromosome and base position. Red diamonds indicate genome-wide significant independent hits (within a 250Kb window and r^2^ < .1) associations.

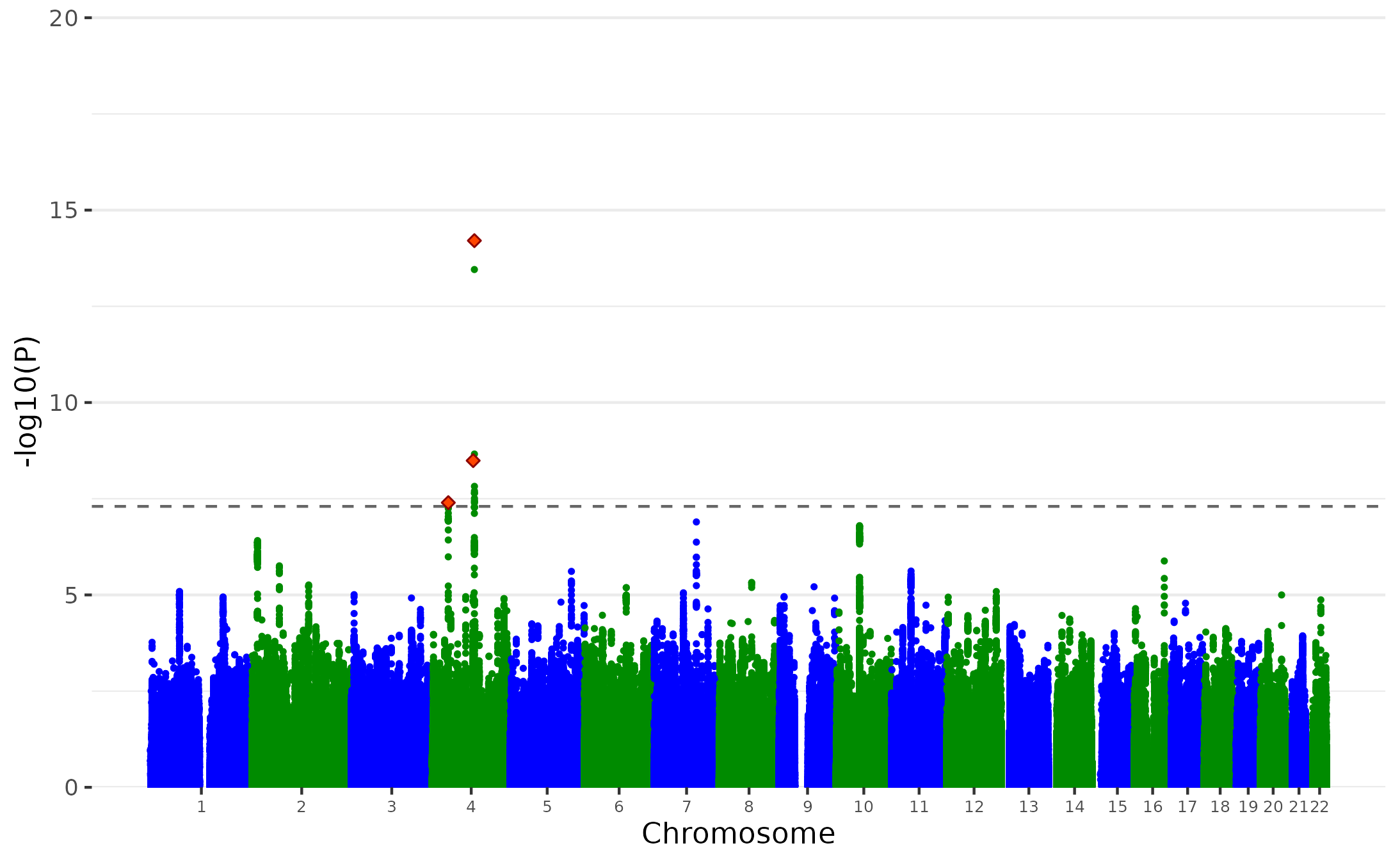

**Supplementary Figure 8. Manhattan plot of the ALCH Non-p GWAS.** Plot of the -log_10_(p-value) associated with the Wald test (two-sided) of β_p_ for all SNPs ordered by chromosome and base position. Red diamonds indicate genome-wide significant independent hits (within a 250Kb window and r^2^ < .1) associations. We identified 2 independent genome-wide significant lead SNPs for ALCH Non-p GWAS.

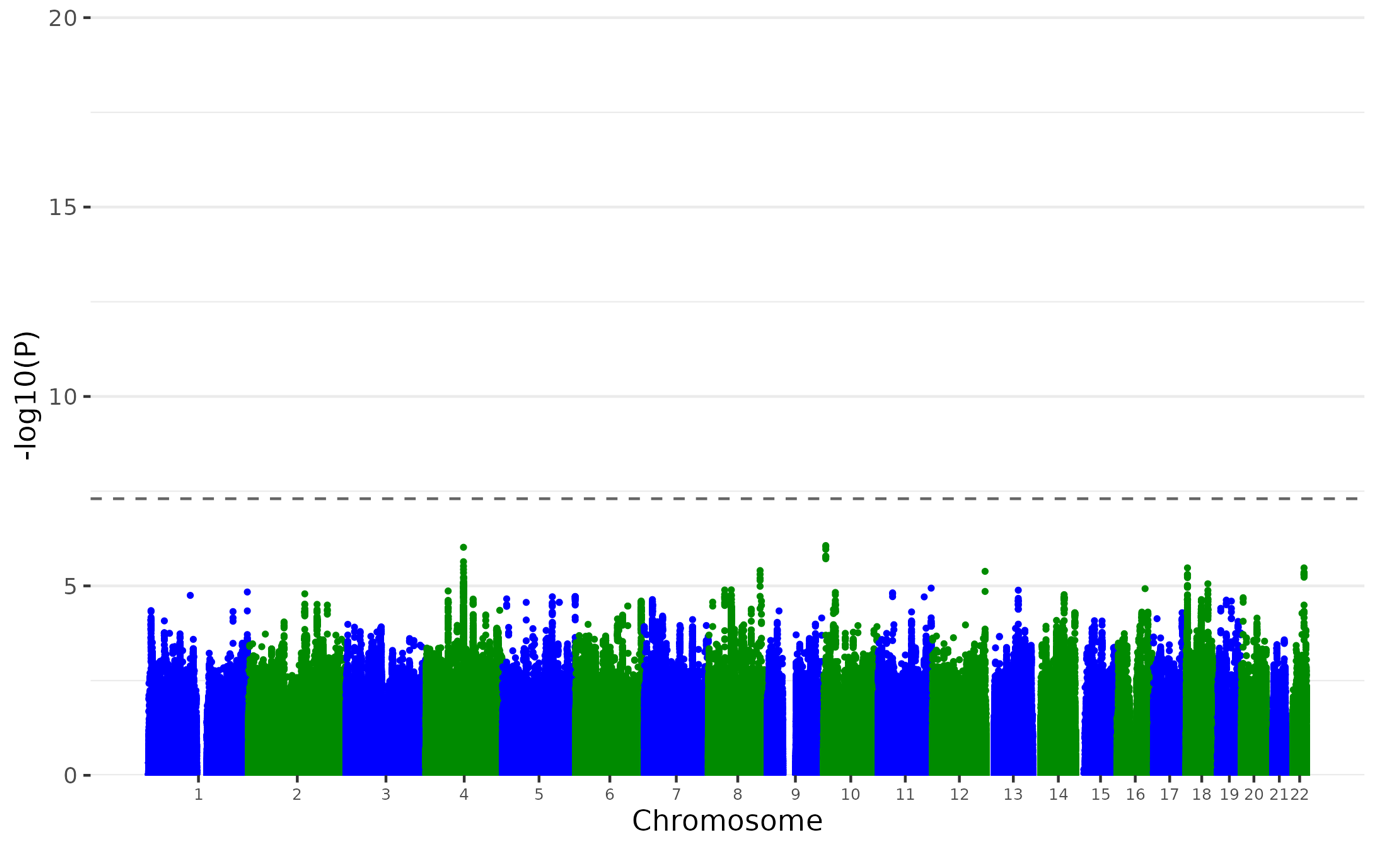

**Supplementary Figure 9. Manhattan plot of the OCD Non-p GWAS.** Plot of the -log_10_(p-value) associated with the Wald test (two-sided) of β_p_ for all SNPs ordered by chromosome and base position. Red diamonds indicate genome-wide significant independent hits (within a 250Kb window and r^2^ < .1) associations.

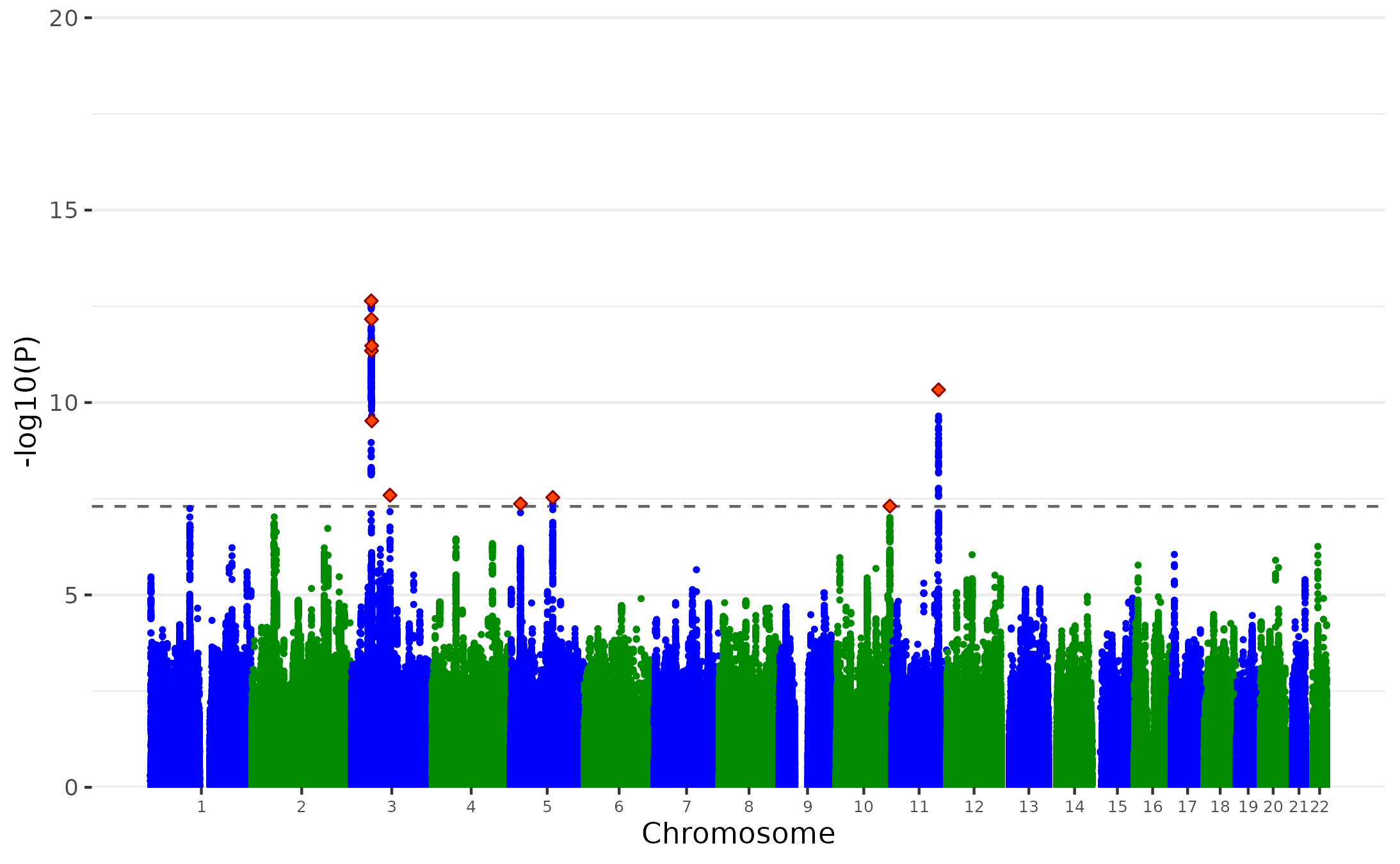

**Supplementary Figure 10. Manhattan plot of the AN Non-p GWAS.** Plot of the -log_10_(p-value) associated with the Wald test (two-sided) of β_p_ for all SNPs ordered by chromosome and base position. Red diamonds indicate genome-wide significant independent hits (within a 250Kb window and r^2^ < .1) associations. We identified 6 independent genome-wide significant lead SNPs for AN Non-p GWAS.

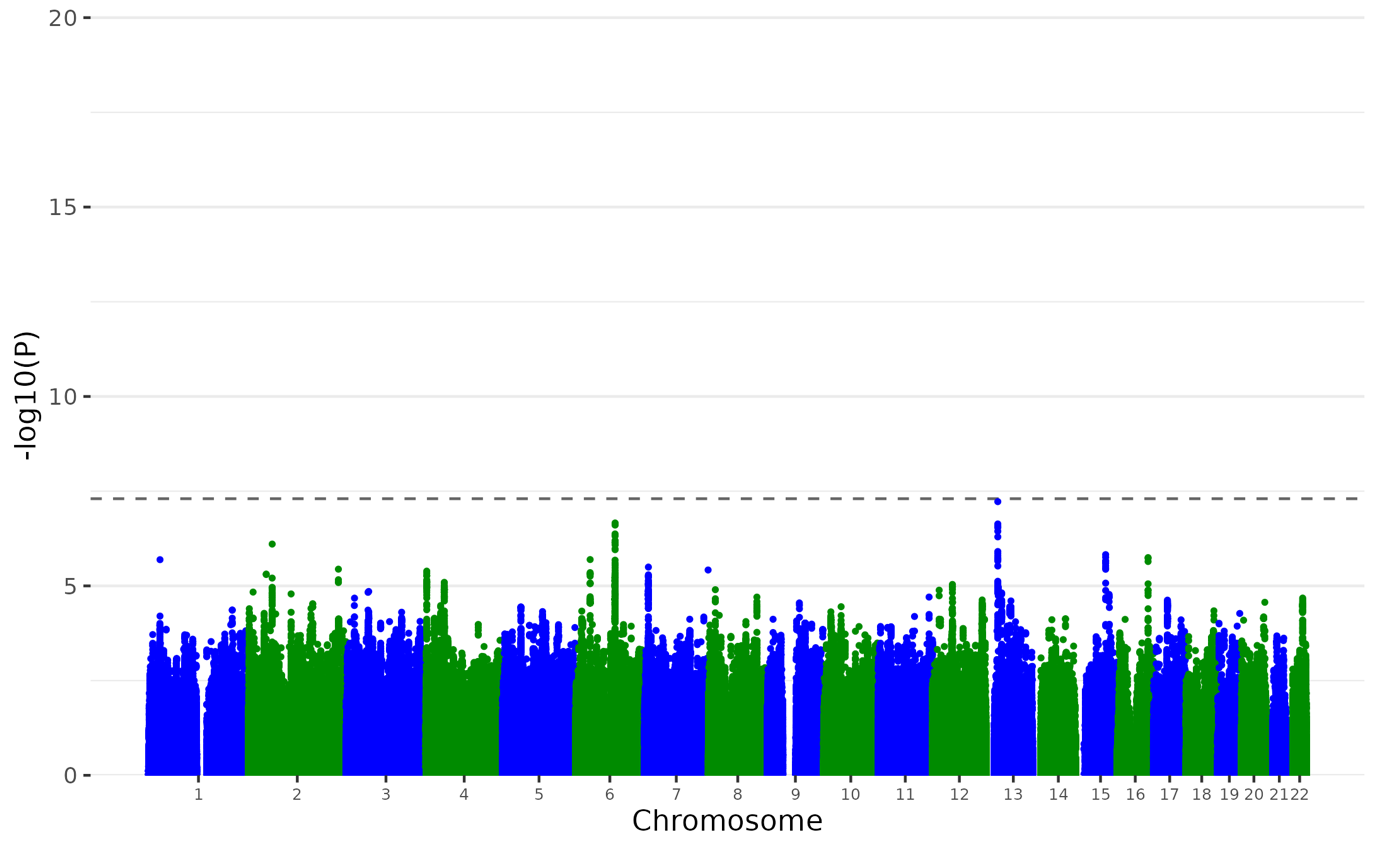

**Supplementary Figure 11. Manhattan plot of the TS Non-p GWAS.** Plot of the -log_10_(p-value) associated with the Wald test (two-sided) of β_p_ for all SNPs ordered by chromosome and base position. Red diamonds indicate genome-wide significant independent hits (within a 250Kb window and r^2^ < .1) associations.

MDD uncorrected for p

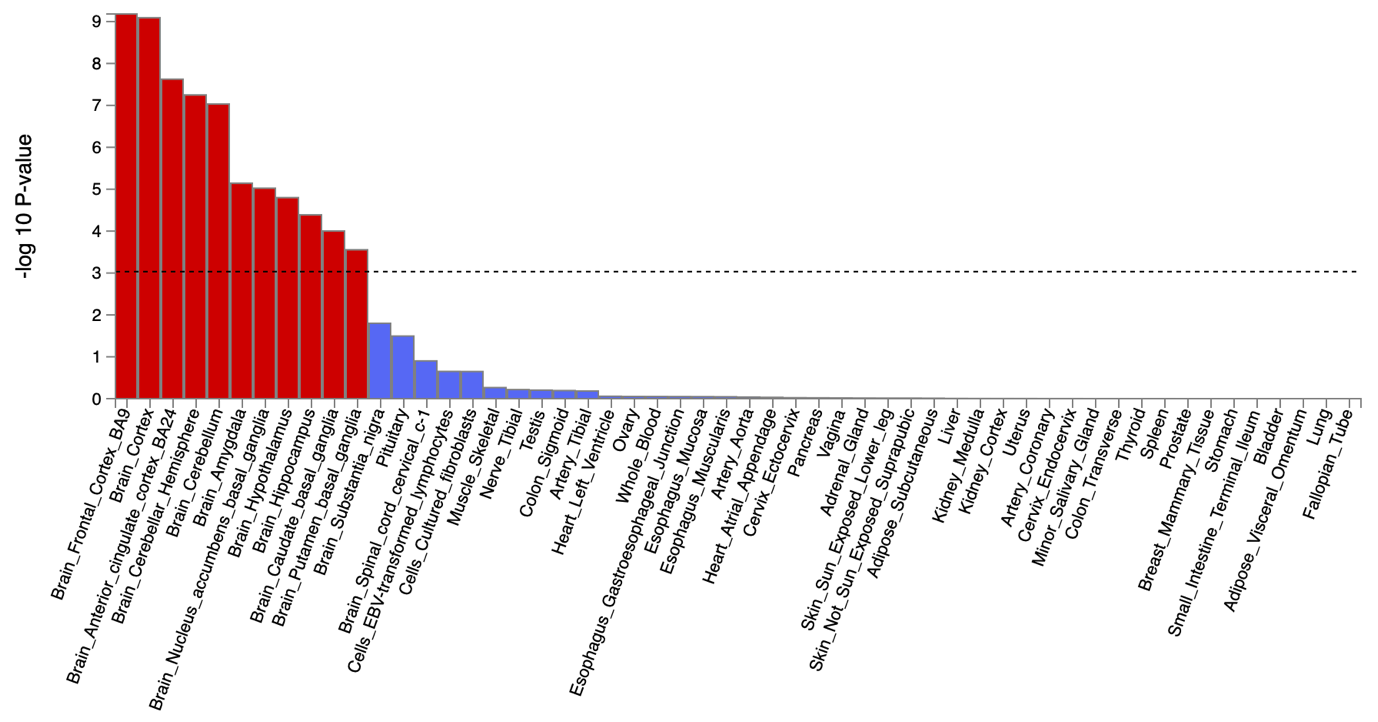

MDD corrected for p

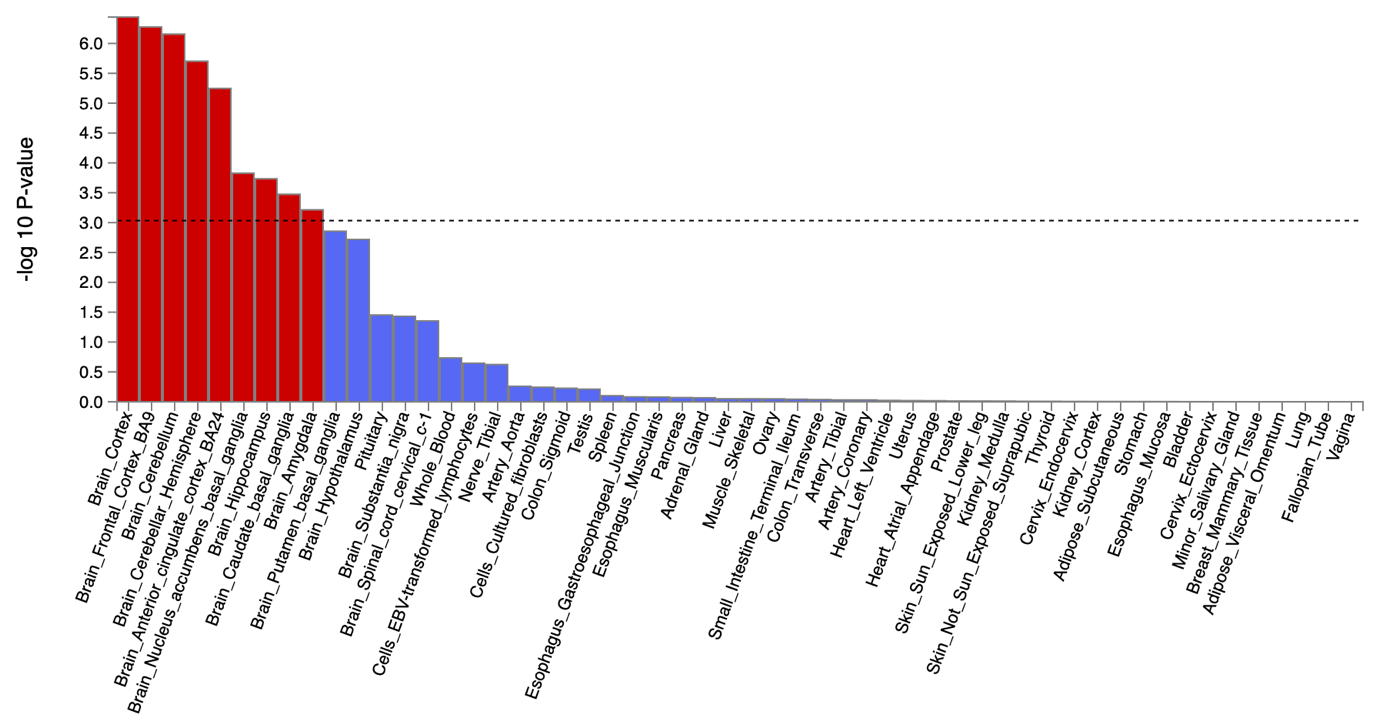

**Supplementary** **Figure 12. The tissue type enrichment results on 53 specific tissue types by GTEx of MDD uncorrected and corrected for p**

Results (-log10 (one-sided P-value)) from MAGMA gene-property analysis of relationships between tissue specific gene expression profiles (GTEx v.7) and Bipolar associations before and after correcting for p. The test was performed for average gene-expression per. tissue type conditioning on the average expression across all categories. Dotted lines indicate significant results after Bonferroni correction. The full results are available in Supplementary Tables 22-25.

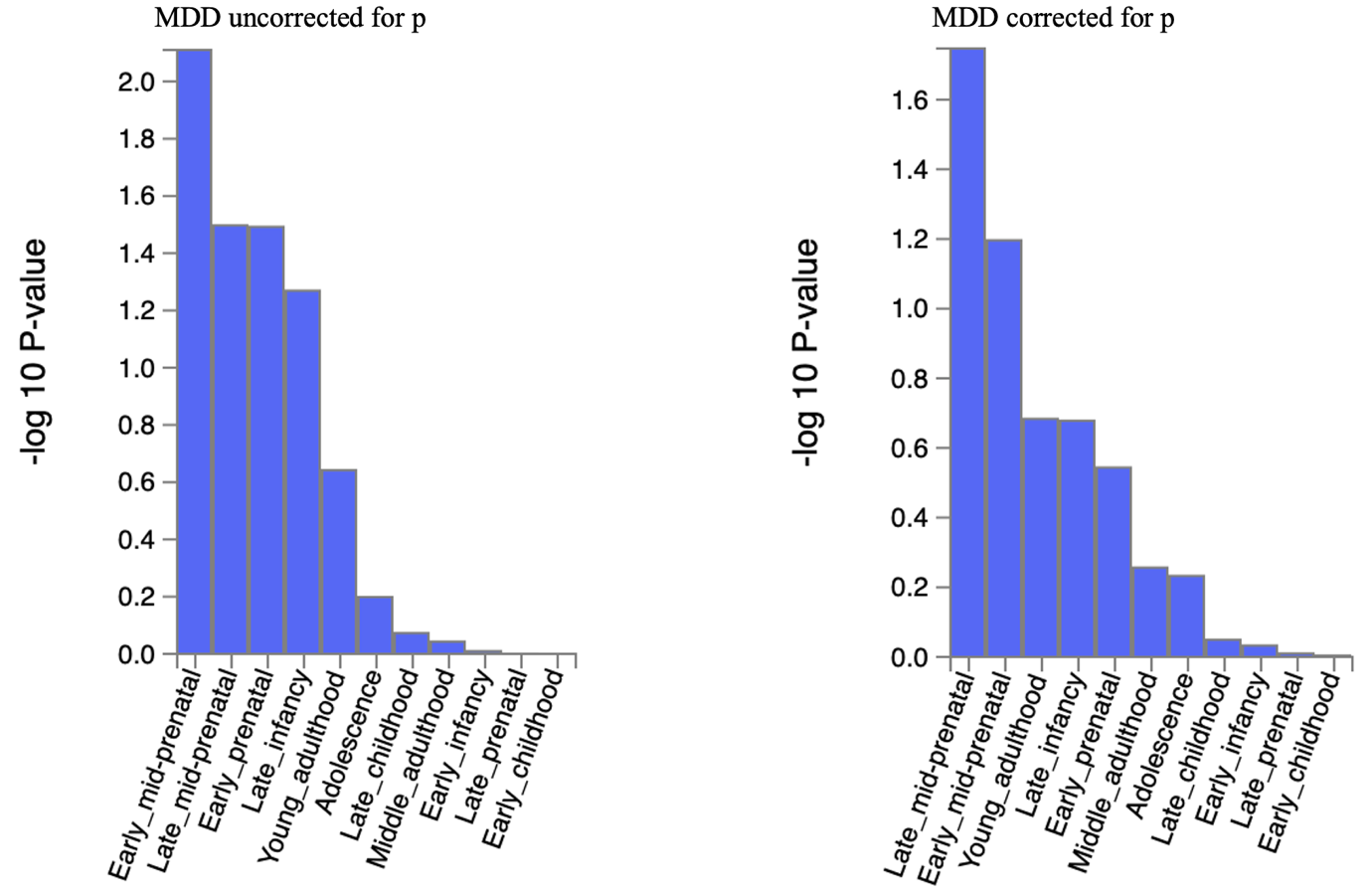

**Supplementary** **Figure 13. The brain sample enrichment results on 11 general brain developmental stages by BrainSpain of MDD uncorrected and corrected for p**

Results (-log10 (one-sided P-value)) from MAGMA gene-property analysis of relationships between gene expression data of developmental brain samples (BrainSpan) and Bipolar associations before and after correcting for p. The test was performed for average gene-expression per. brain sample conditioning on the average expression across all developmental stages. Dotted lines indicate significant results after Bonferroni correction. The full results are available in Supplementary Tables 22-25.

BIP uncorrected for p

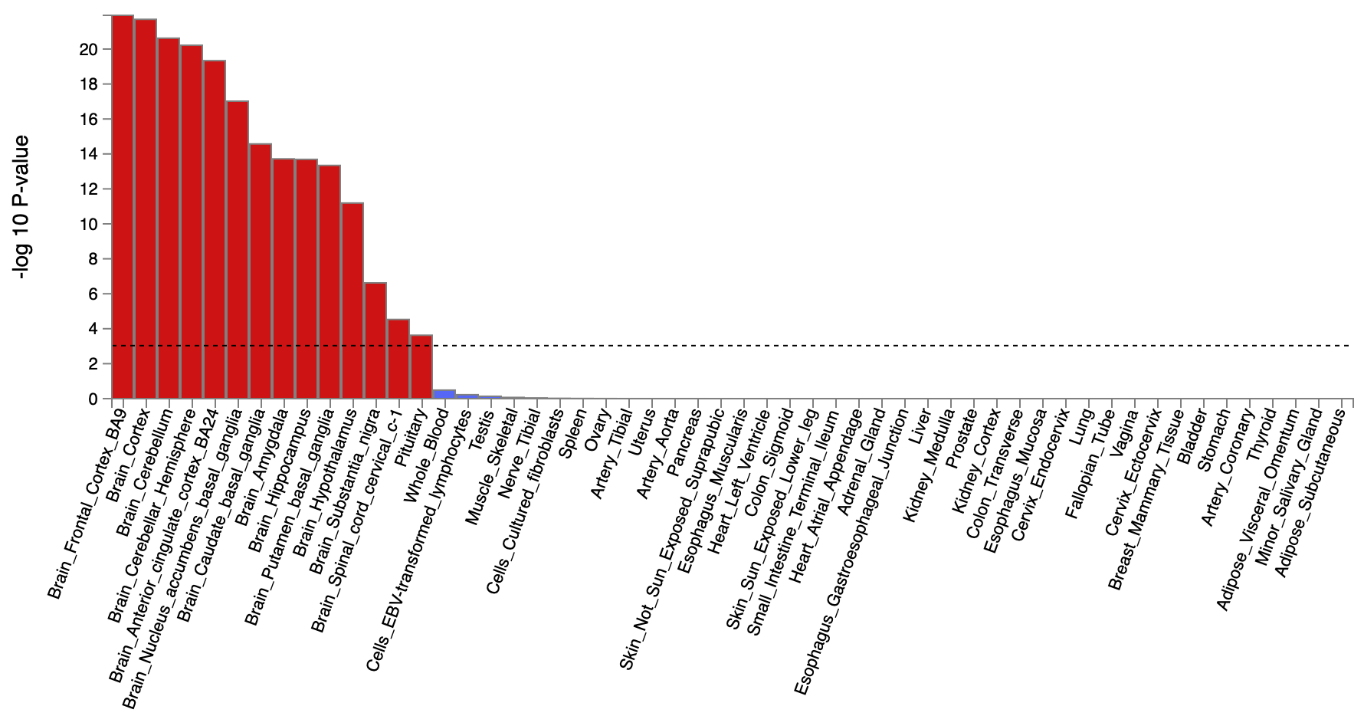

BIP corrected for p

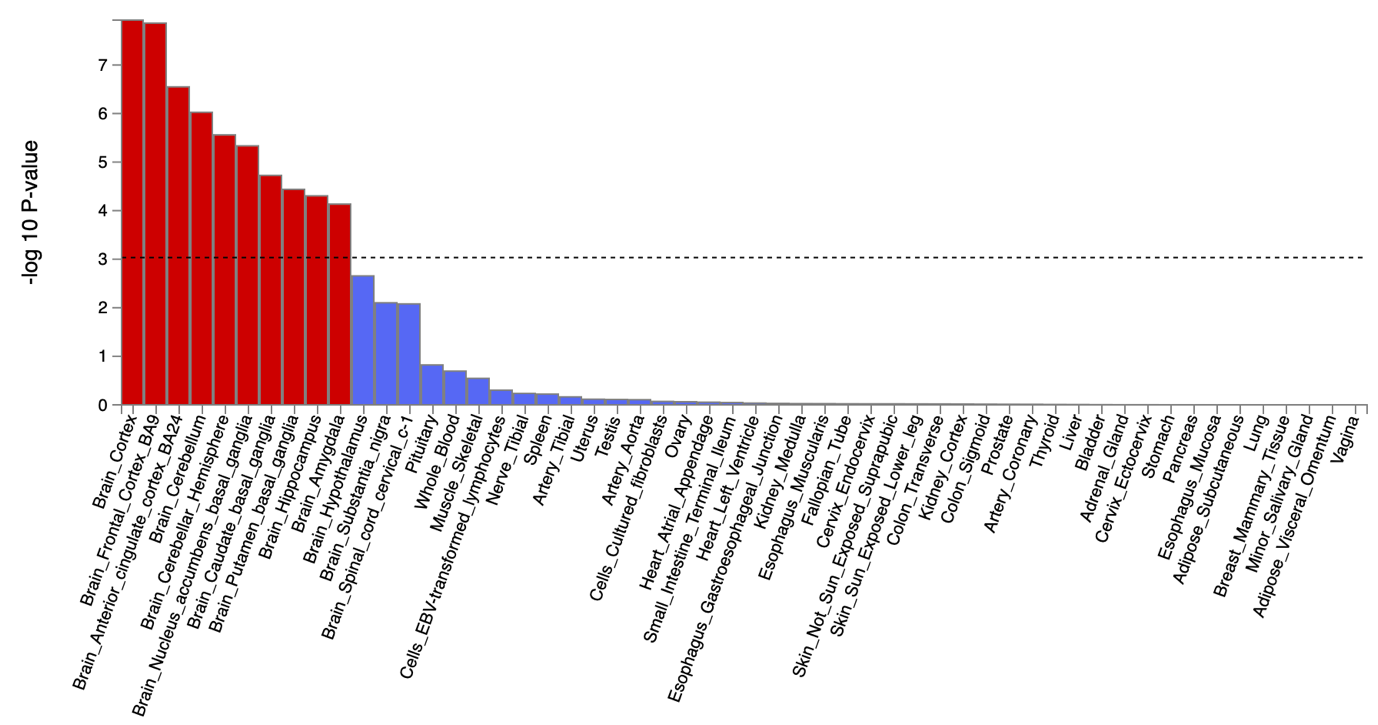

**Supplementary** **Figure 14. The tissue type enrichment results on 53 specific tissue types by GTEx of BIP uncorrected and corrected for p**

Results (-log10 (one-sided P-value)) from MAGMA gene-property analysis of relationships between tissue specific gene expression profiles (GTEx v.7) and Bipolar associations before and after correcting for p. The test was performed for average gene-expression per. tissue type conditioning on the average expression across all categories. Dotted lines indicate significant results after Bonferroni correction. The full results are available in Supplementary Tables 22-25.

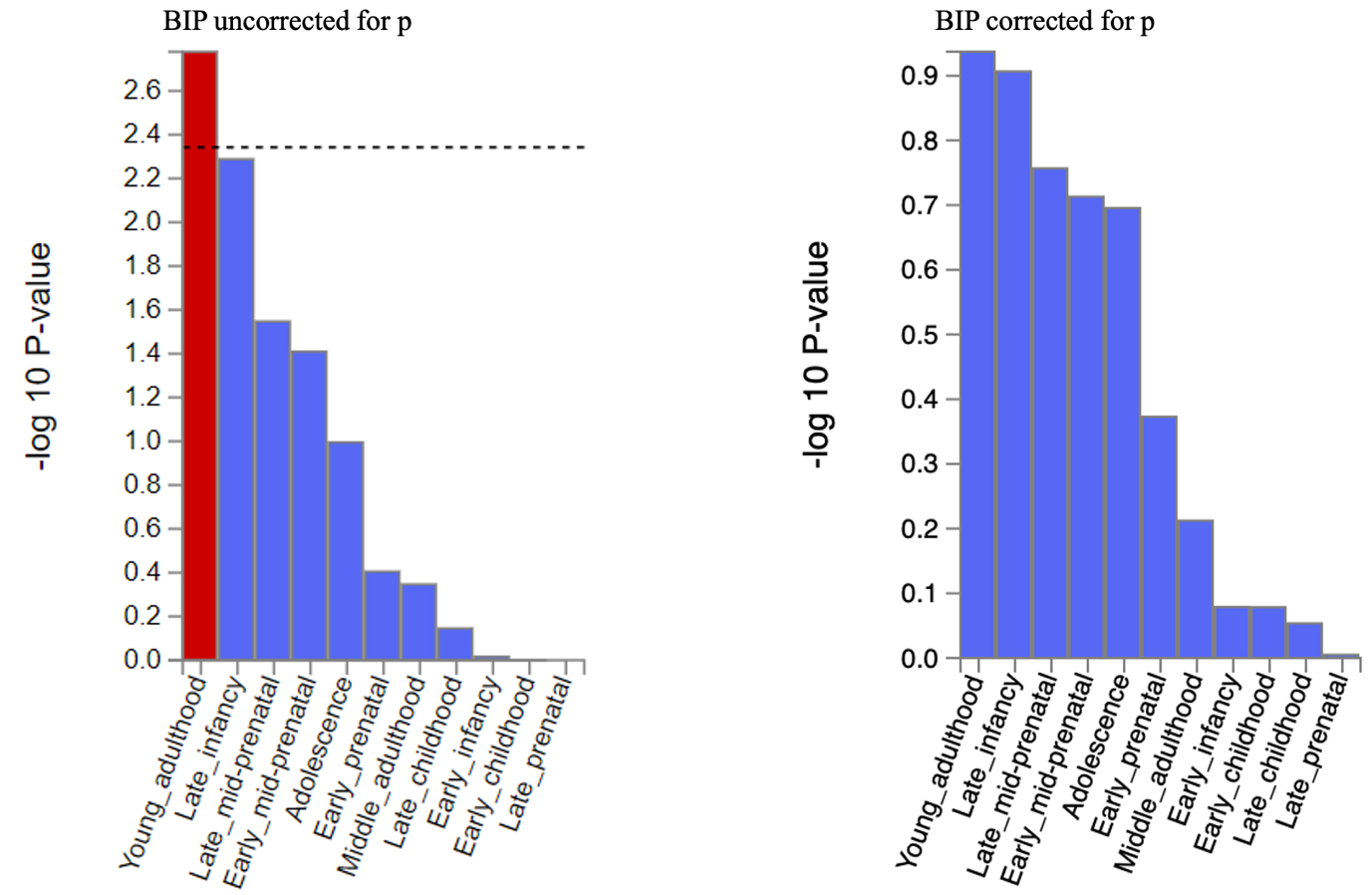

**Supplementary** **Figure 15. The brain sample enrichment results on 11 general brain developmental stages by BrainSpain of Bipolar uncorrected and corrected for p**

Results (-log10 (one-sided P-value)) from MAGMA gene-property analysis of relationships between gene expression data of developmental brain samples (BrainSpan) and Bipolar associations before and after correcting for p. The test was performed for average gene-expression per. brain sample conditioning on the average expression across all developmental stages. Dotted lines indicate significant results after Bonferroni correction. The full results are available in Supplementary Tables 22-25.

SCZ uncorrected for p

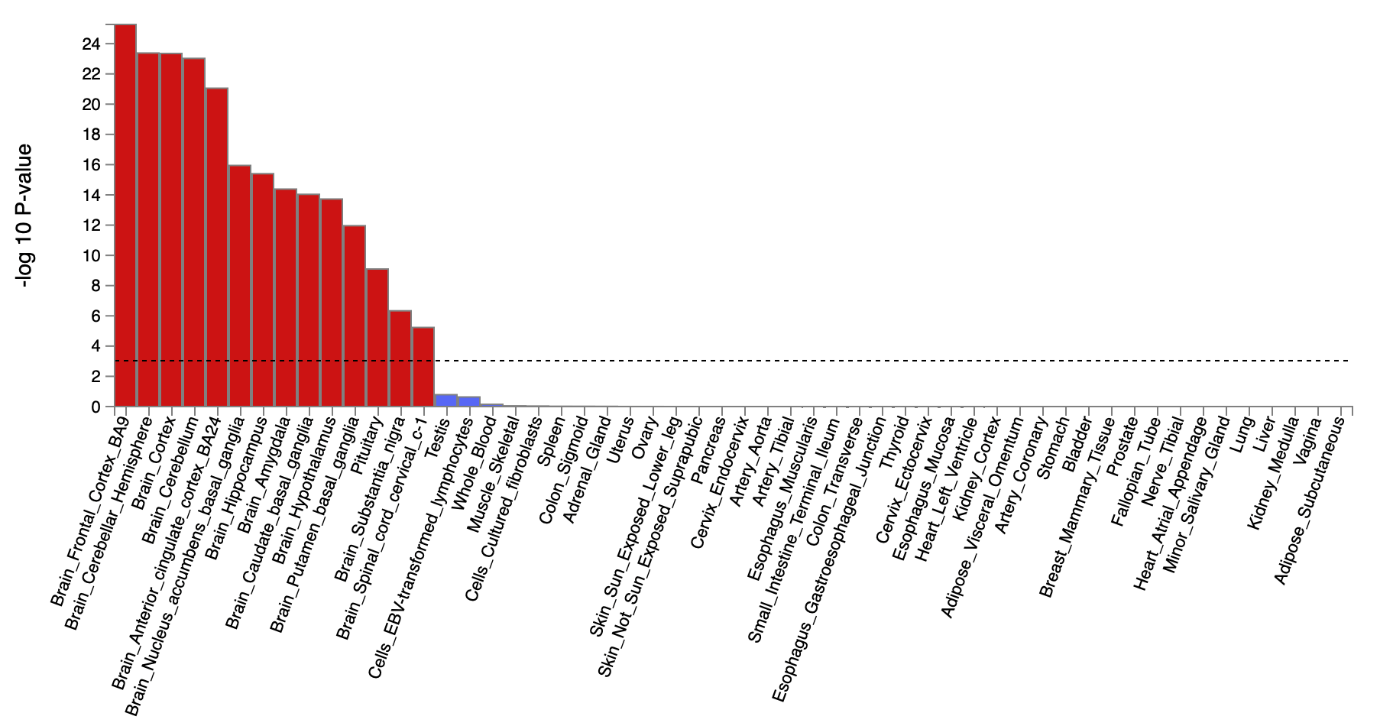

SCZ corrected for p

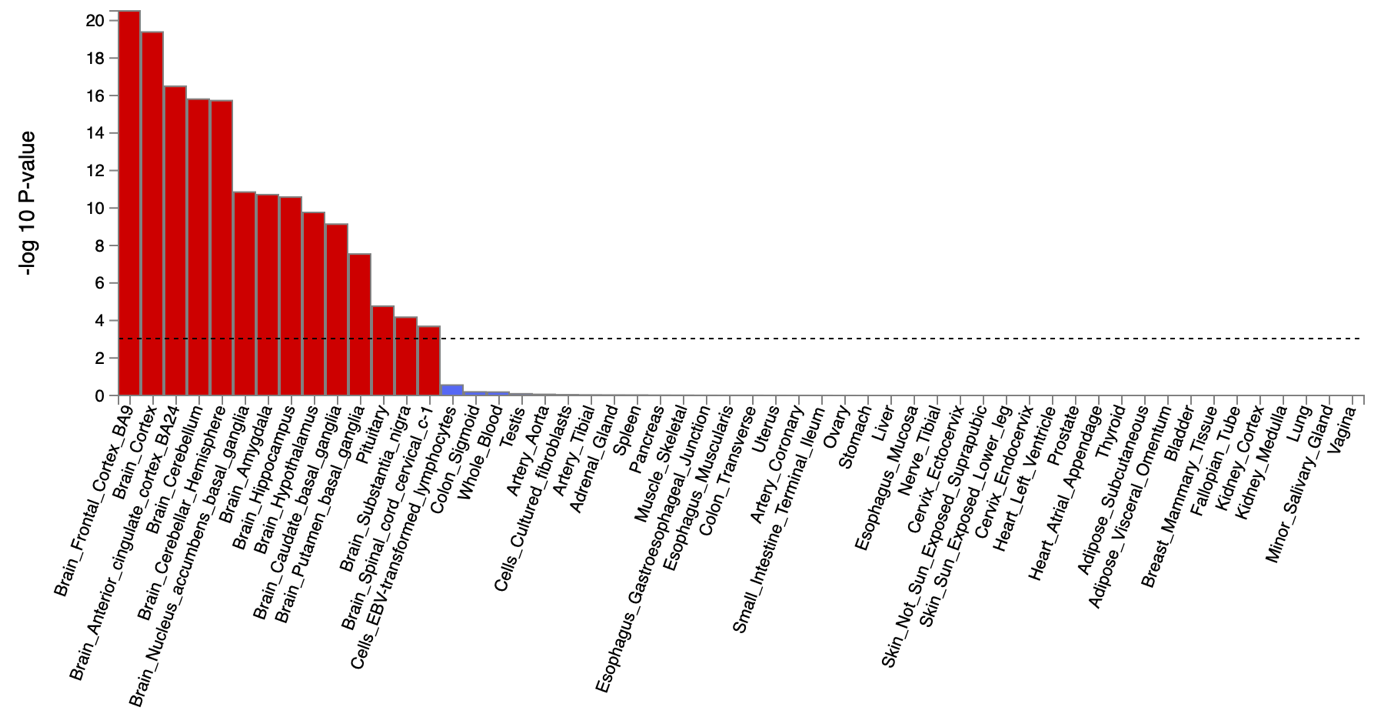

**Supplementary** **Figure 16. The tissue type enrichment results on 53 specific tissue types by GTEx of Schizophrenia uncorrected and corrected for p**

Results (-log10 (one-sided P-value)) from MAGMA gene-property analysis of relationships between tissue specific gene expression profiles (GTEx v.7) and schizophrenia associations before and after correcting for p. The test was performed for average gene-expression per tissue type conditioning on the average expression across all categories. Dotted lines indicate significant results after Bonferroni correction. The full results are available in Supplementary Tables 18-21.

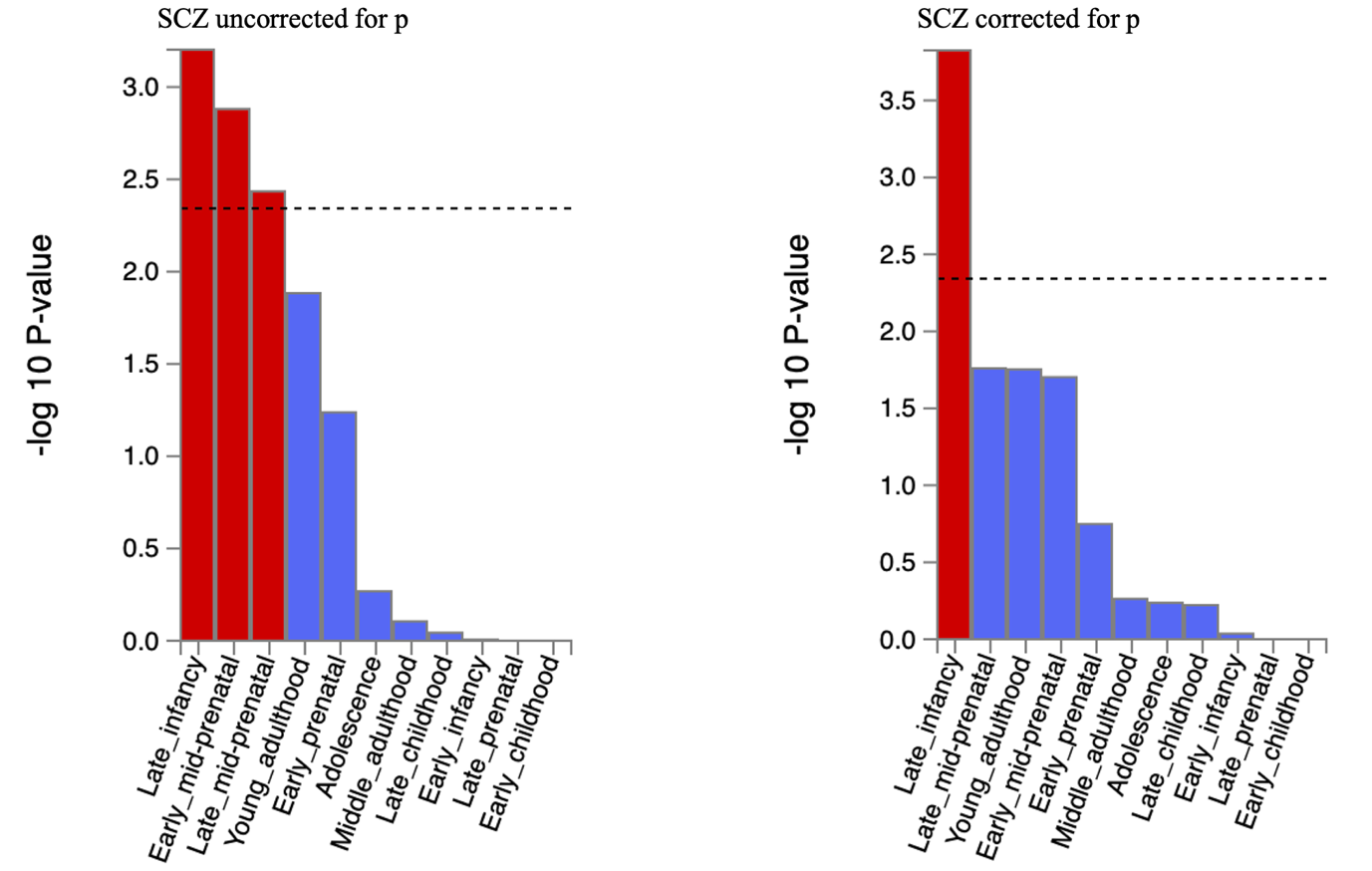

**Supplementary** **Figure 17. The brain sample enrichment results for 11 general brain developmental stages by BrainSpan for schizophrenia uncorrected and corrected for p**

Results (-log10 (one-sided P-value)) from MAGMA gene-property analysis of relationships between gene expression data of developmental brain samples (BrainSpan) and schizophrenia associations before and after correcting for p. The test was performed for average gene-expression per brain sample conditioning on the average expression across all developmental stages. Dotted lines indicate significant results after Bonferroni correction. The full results are available in Supplementary Tables 18-21.

ADHD uncorrected for p

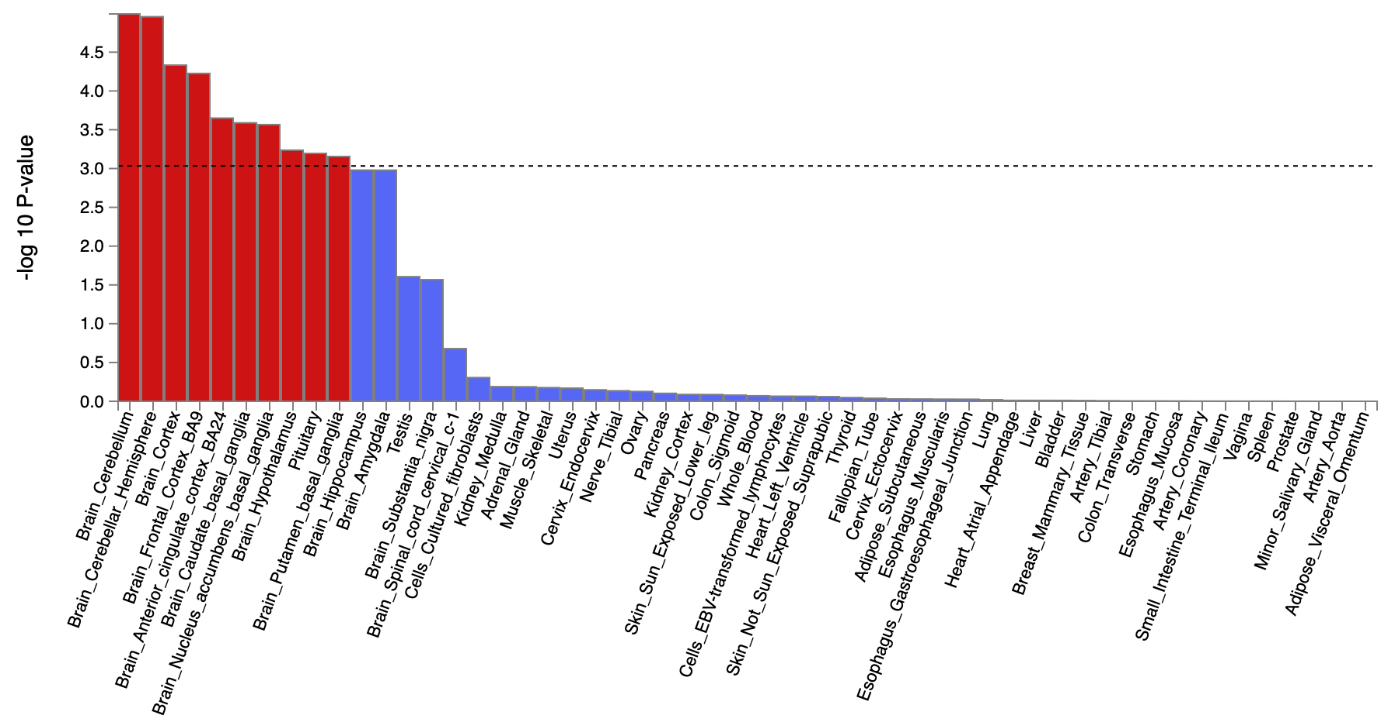

ADHD corrected for p

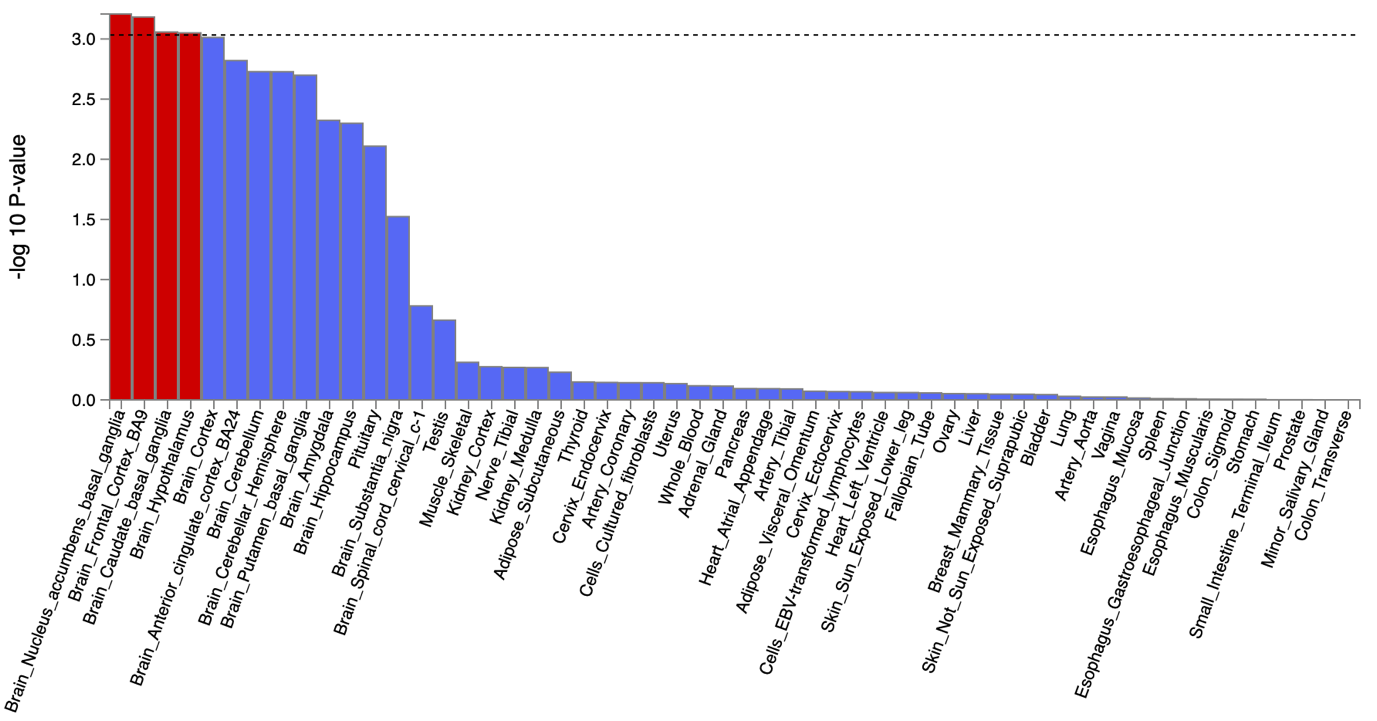

**Supplementary** **Figure 18. The tissue type enrichment results on 53 specific tissue types by GTEx of ADHD uncorrected and corrected for p**

Results (-log10 (one-sided P-value)) from MAGMA gene-property analysis of relationships between tissue specific gene expression profiles (GTEx v.7) and ADHD associations before and after correcting for p. The test was performed for average gene-expression per. tissue type conditioning on the average expression across all categories. Dotted lines indicate significant results after Bonferroni correction. The full results are available in Supplementary Tables 26-29.

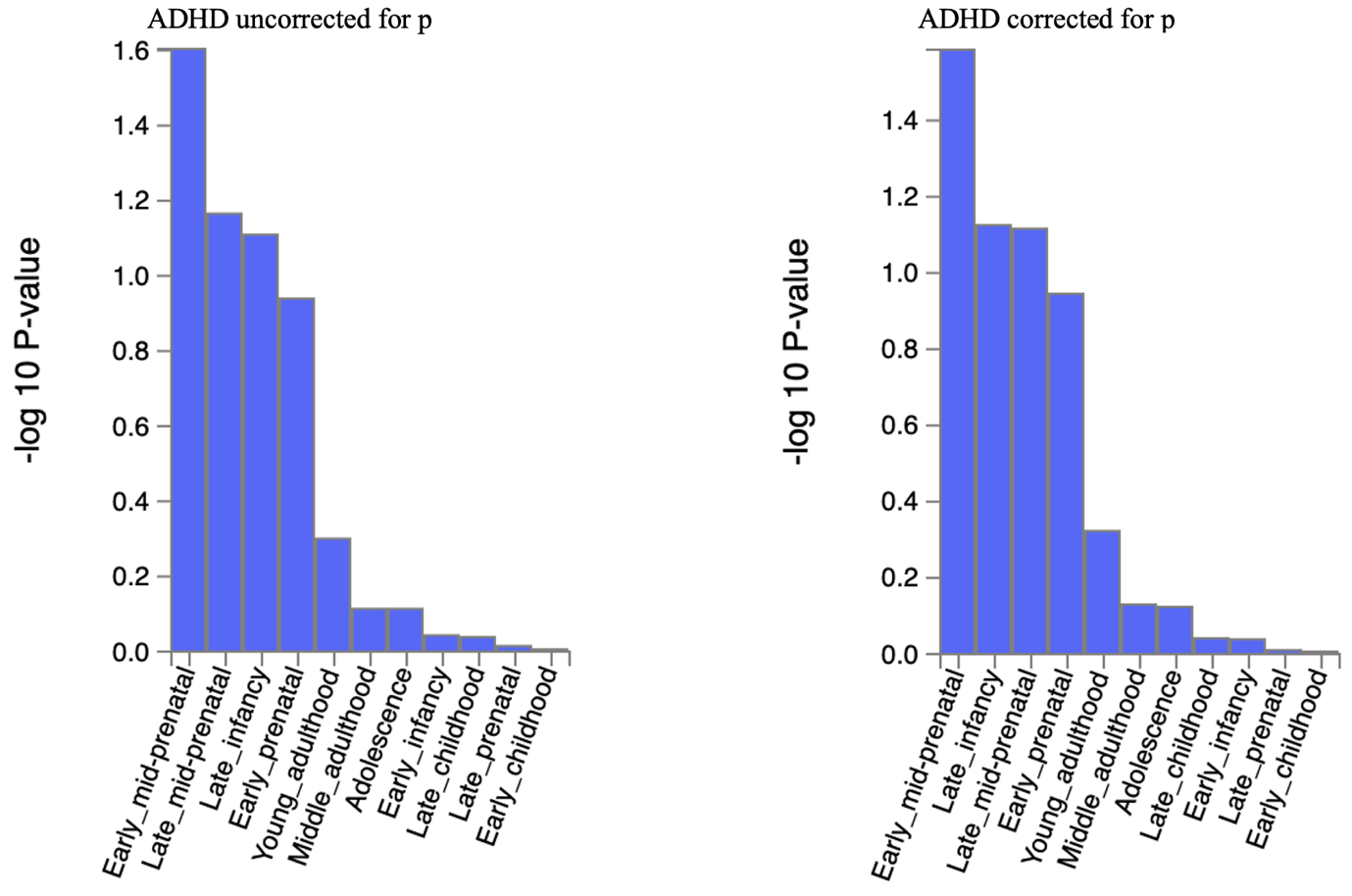

**Supplementary** **Figure 19. The brain sample enrichment results on 11 general brain developmental stages by BrainSpain of ADHD uncorrected and corrected for p**

Results (-log10 (one-sided P-value)) from MAGMA gene-property analysis of relationships between gene expression data of developmental brain samples (BrainSpan) and ADHD associations before and after correcting for p. The test was performed for average gene-expression per. brain sample conditioning on the average expression across all developmental stages. Dotted lines indicate significant results after Bonferroni correction. The full results are available in Supplementary Tables 26-29.

ASD uncorrected for p

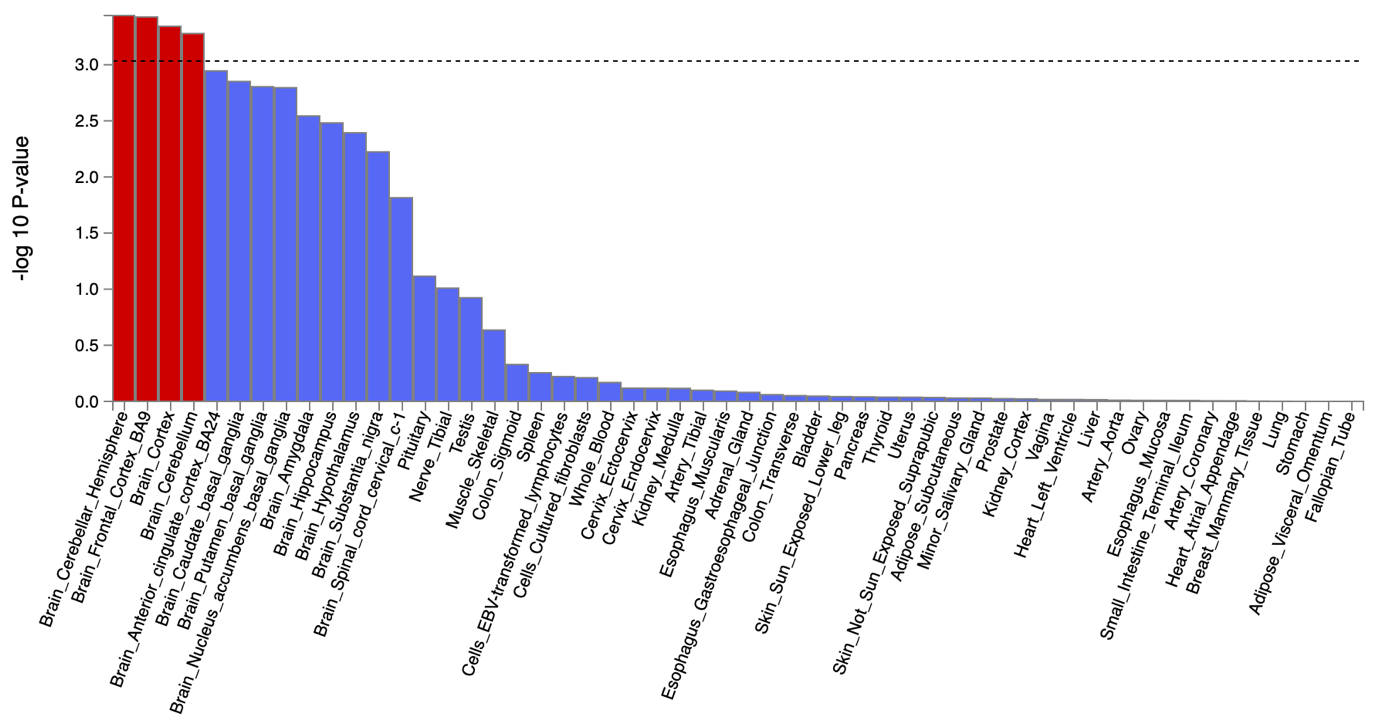

ASD corrected for p

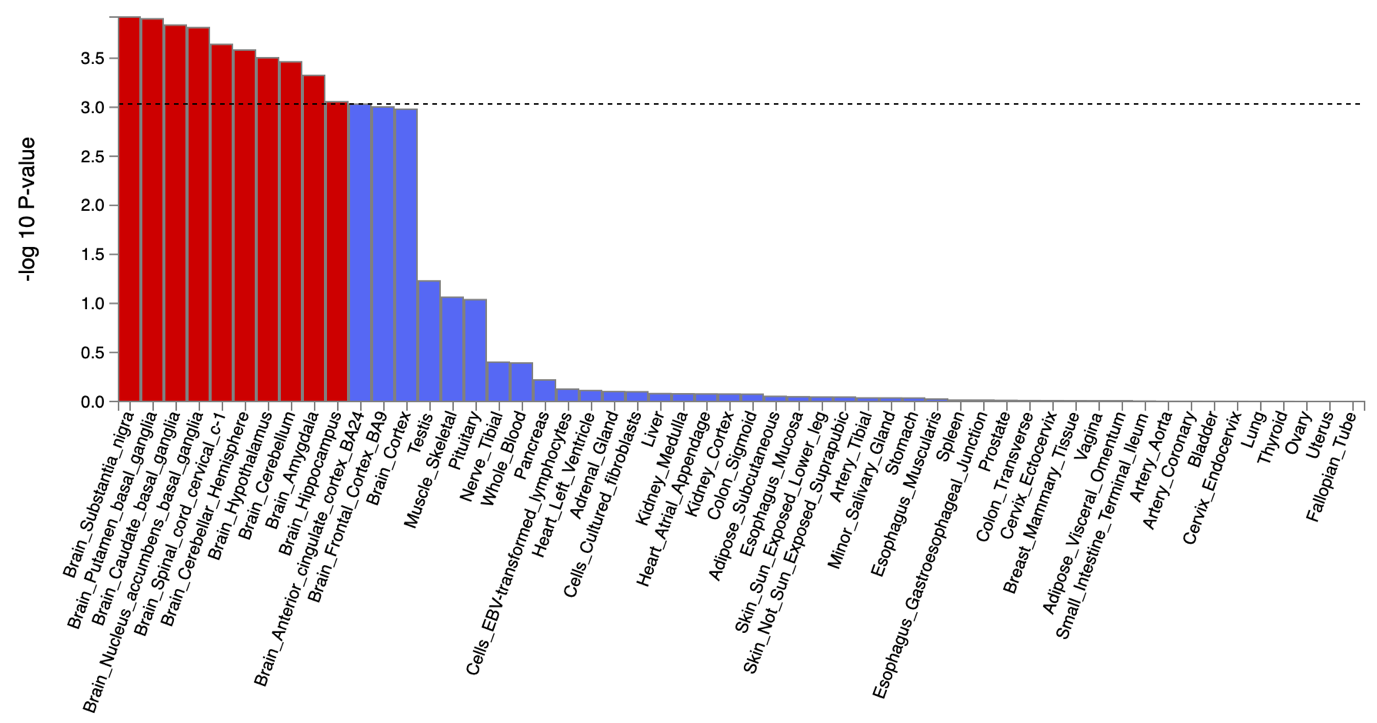

**Supplementary** **Figure 20. The tissue type enrichment results on 53 specific tissue types by GTEx of ASD uncorrected and corrected for p**

Results (-log10 (one-sided P-value)) from MAGMA gene-property analysis of relationships between tissue specific gene expression profiles (GTEx v.7) and ALCH associations before and after correcting for p. The test was performed for average gene-expression per. tissue type conditioning on the average expression across all categories. The full results are available in Supplementary Tables 34-37.

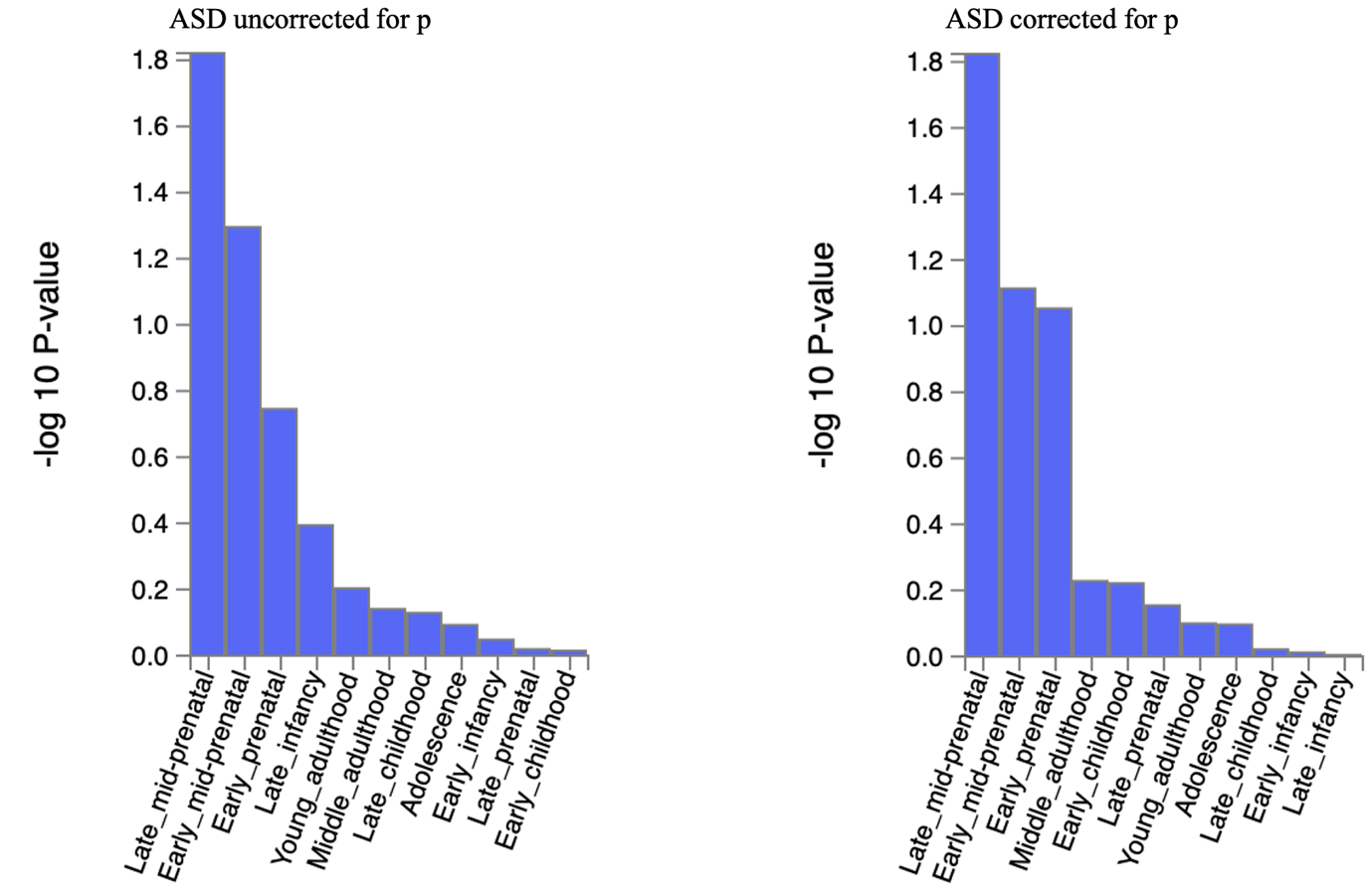

**Supplementary** **Figure 21. The brain sample enrichment results on 11 general brain developmental stages by BrainSpain of ASD uncorrected and corrected for p**

Results (-log10 (one-sided P-value)) from MAGMA gene-property analysis of relationships between gene expression data of developmental brain samples (BrainSpan) and ALCH associations before and after correcting for p. The test was performed for average gene-expression per. brain sample conditioning on the average expression across all developmental stages. Dotted line indicates significant results after Bonferroni correction. The full results are available in Supplementary Tables 34-3

ALCH uncorrected for p

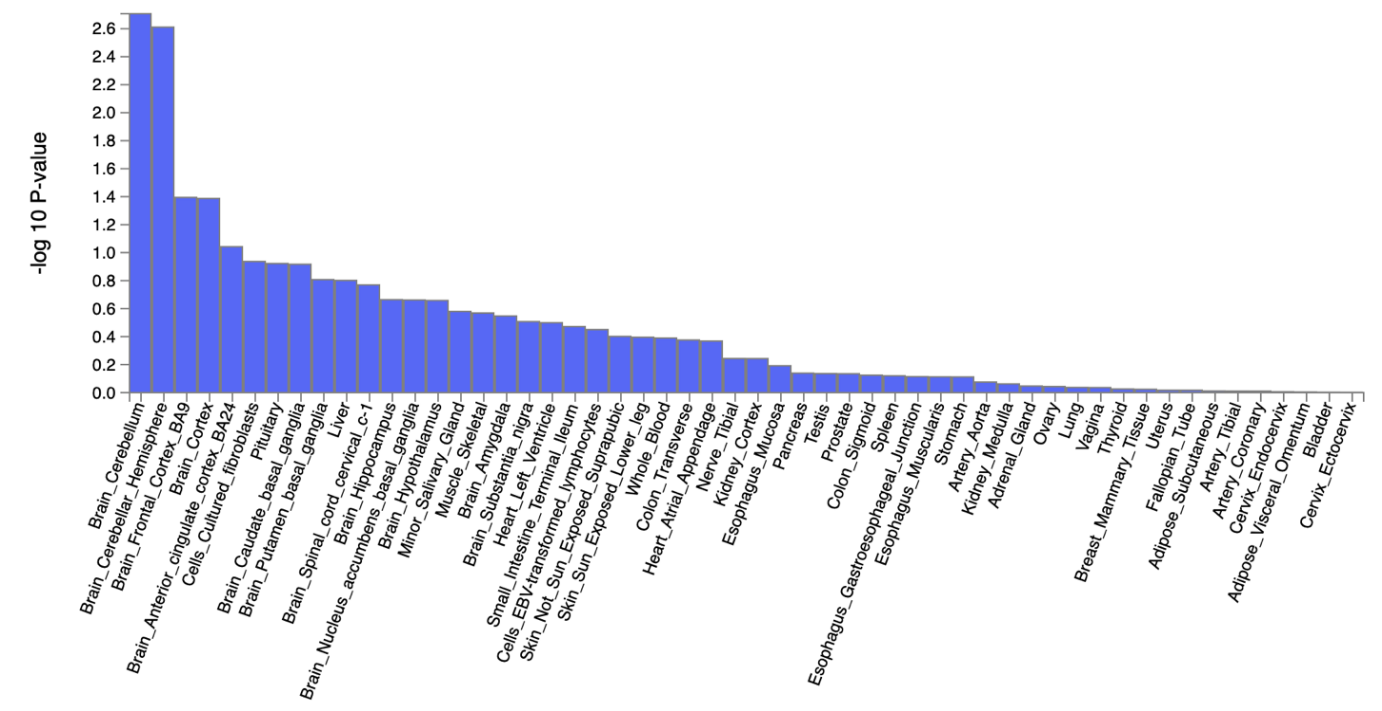

ALCH corrected for p

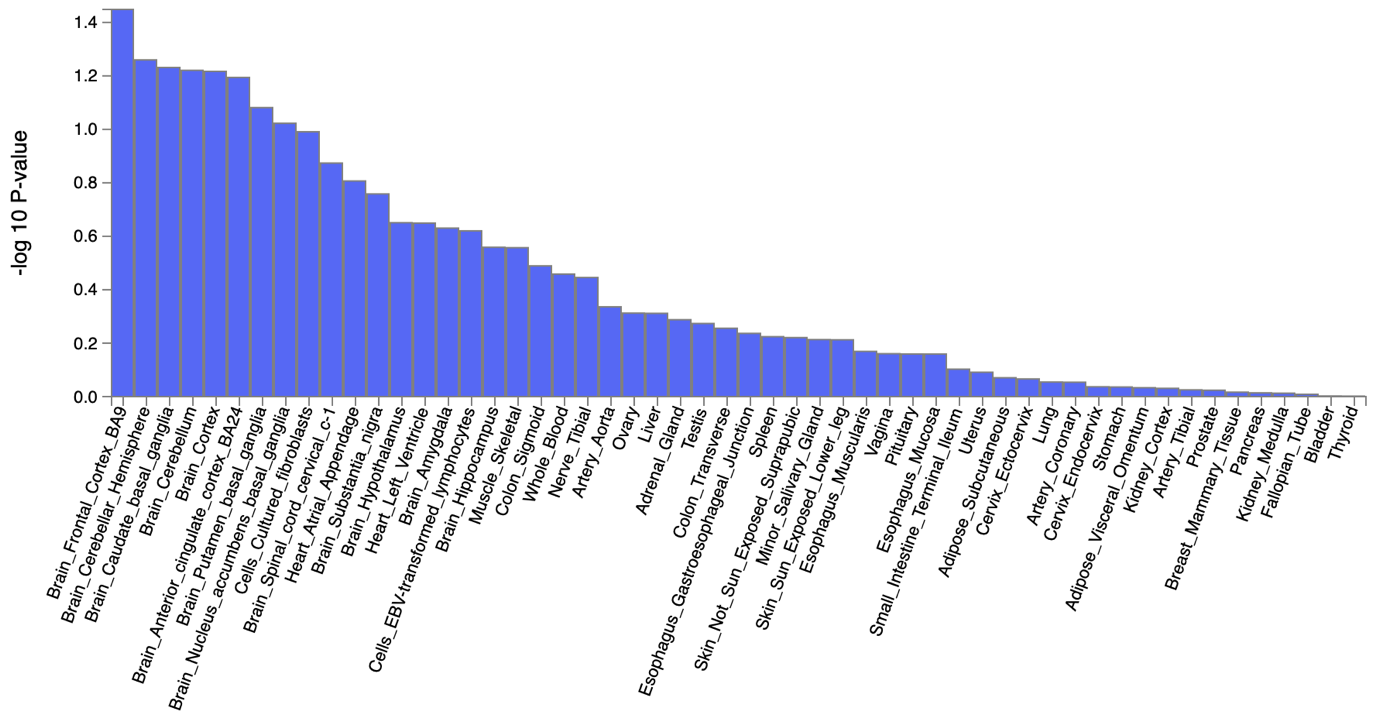

**Supplementary** **Figure 22. The tissue type enrichment results on 53 specific tissue types by GTEx of ALCH uncorrected and corrected for p**

Results (-log10 (one-sided P-value)) from MAGMA gene-property analysis of relationships between tissue specific gene expression profiles (GTEx v.7) and ALCH associations before and after correcting for p. The test was performed for average gene-expression per. tissue type conditioning on the average expression across all categories. The full results are available in Supplementary Tables 34-37.

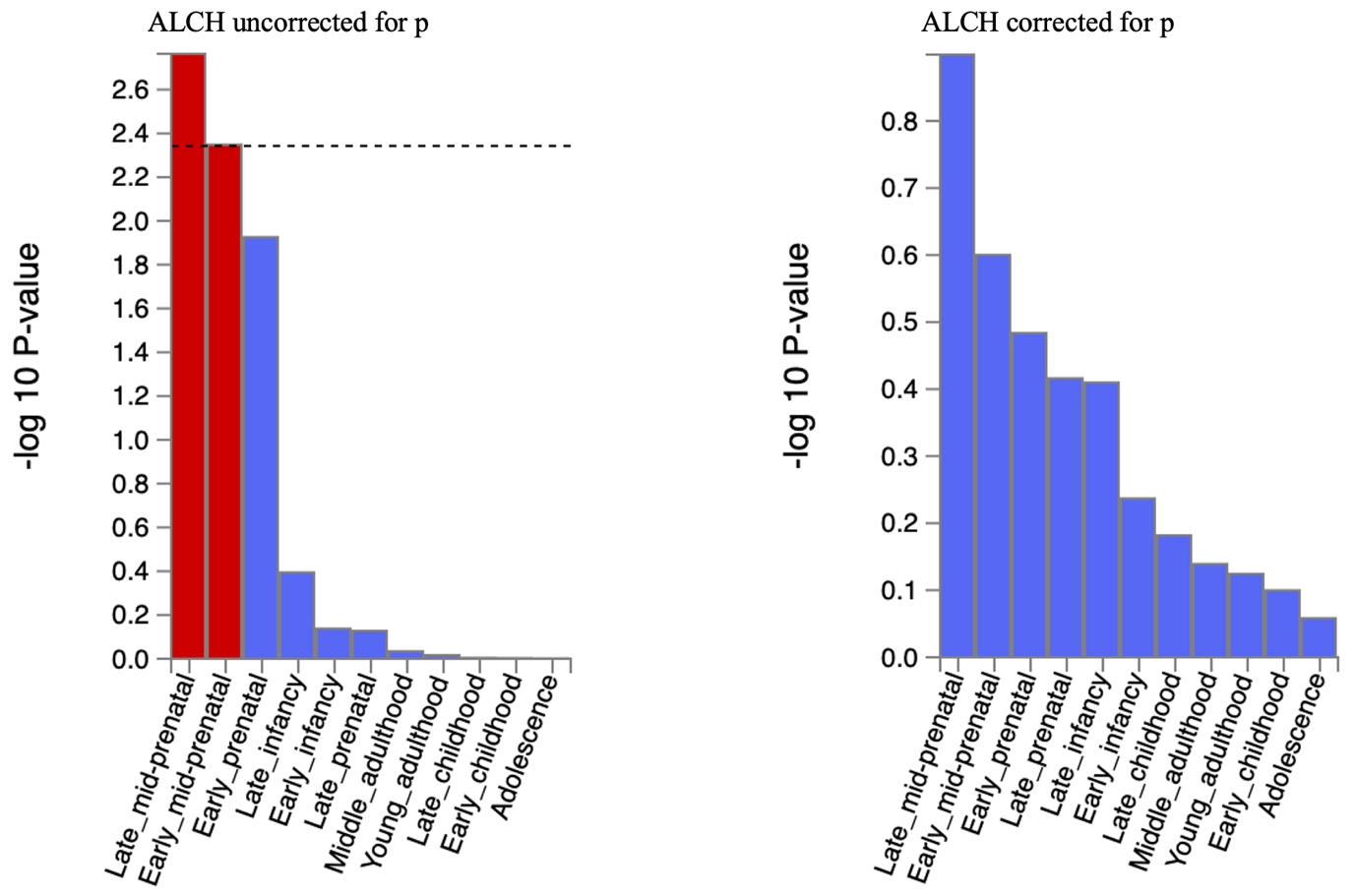

**Supplementary** **Figure 23. The brain sample enrichment results on 11 general brain developmental stages by BrainSpain of ALCH uncorrected and corrected for p**

Results (-log10 (one-sided P-value)) from MAGMA gene-property analysis of relationships between gene expression data of developmental brain samples (BrainSpan) and ALCH associations before and after correcting for p. The test was performed for average gene-expression per. brain sample conditioning on the average expression across all developmental stages. Dotted line indicates significant results after Bonferroni correction. The full results are available in Supplementary Tables 34-37

AN uncorrected for p

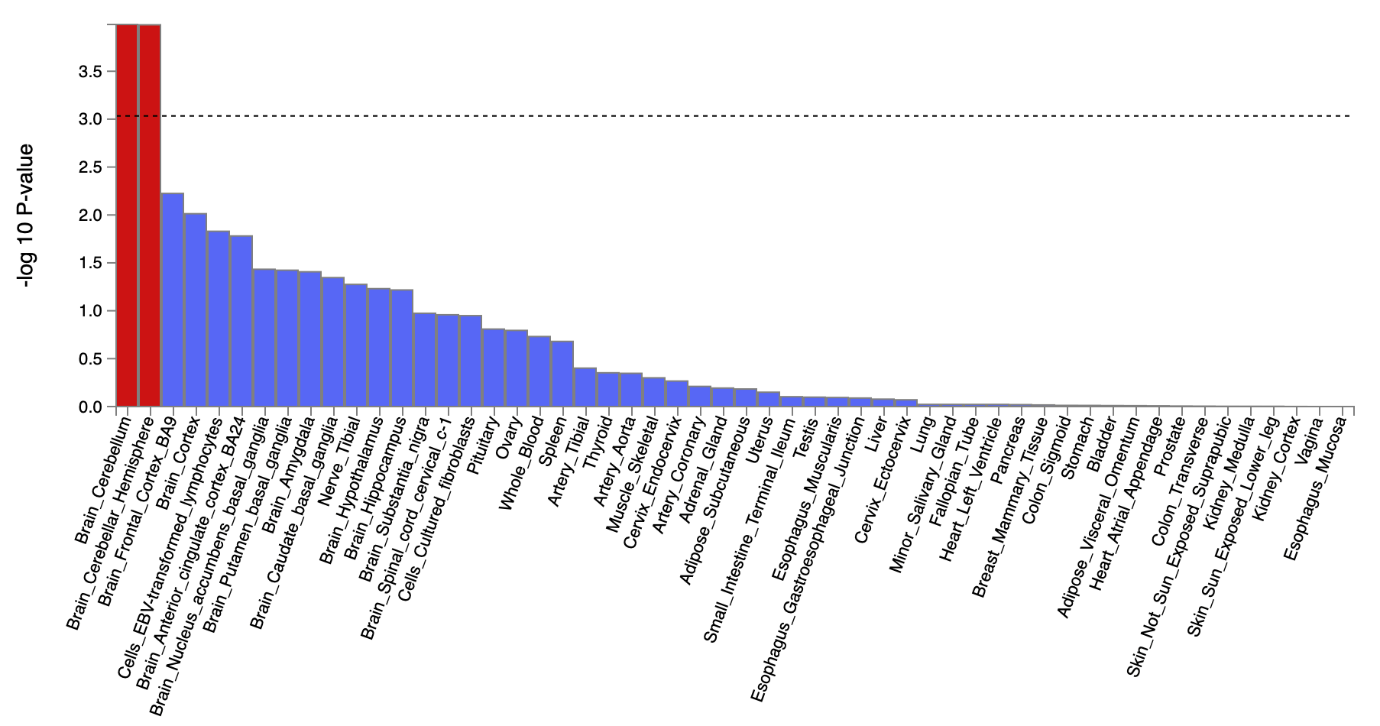

AN corrected for p

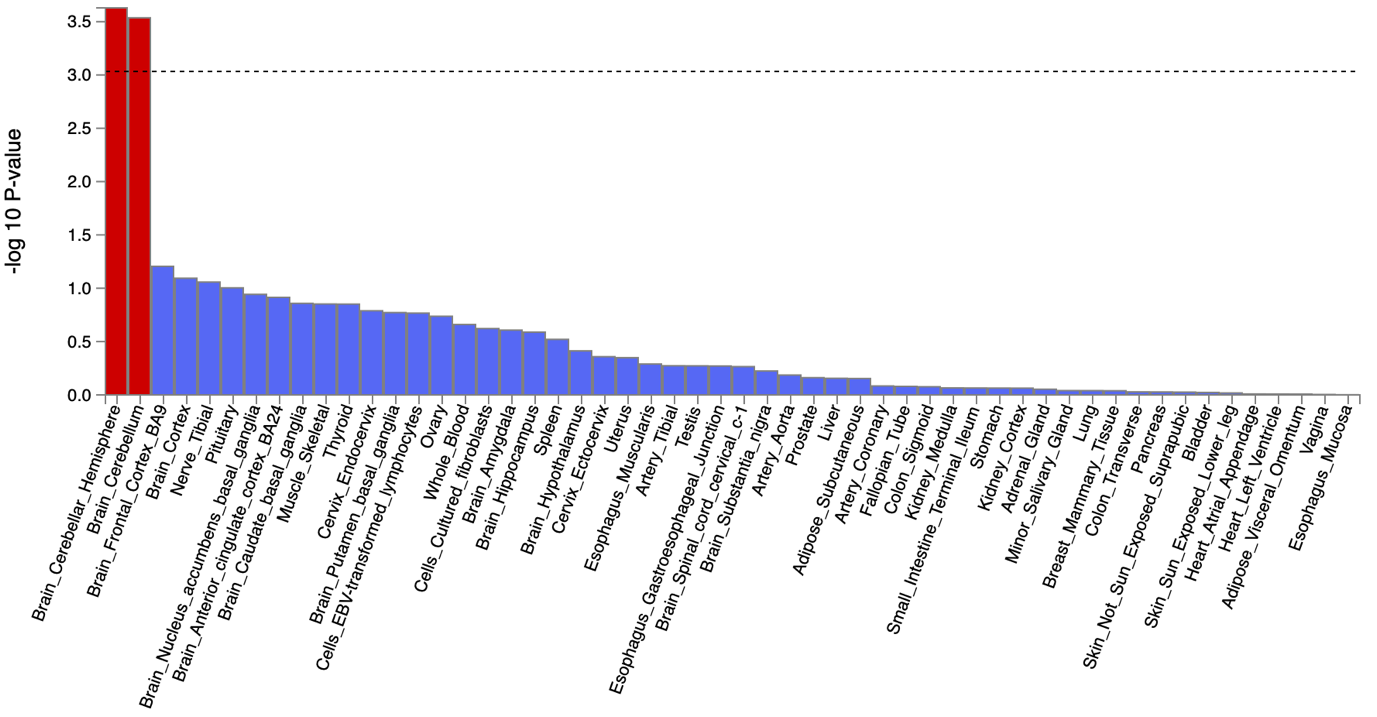

**Supplementary** **Figure 24. The tissue type enrichment results on 53 specific tissue types by GTEx of AN uncorrected and corrected for p**

Results (-log10 (one-sided P-value)) from MAGMA gene-property analysis of relationships between tissue specific gene expression profiles (GTEx v.7) and AN associations before and after correcting for p. The test was performed for average gene-expression per. tissue type conditioning on the average expression across all categories. Dotted lines indicate significant results after Bonferroni correction. The full results are available in Supplementary Tables 30-33.

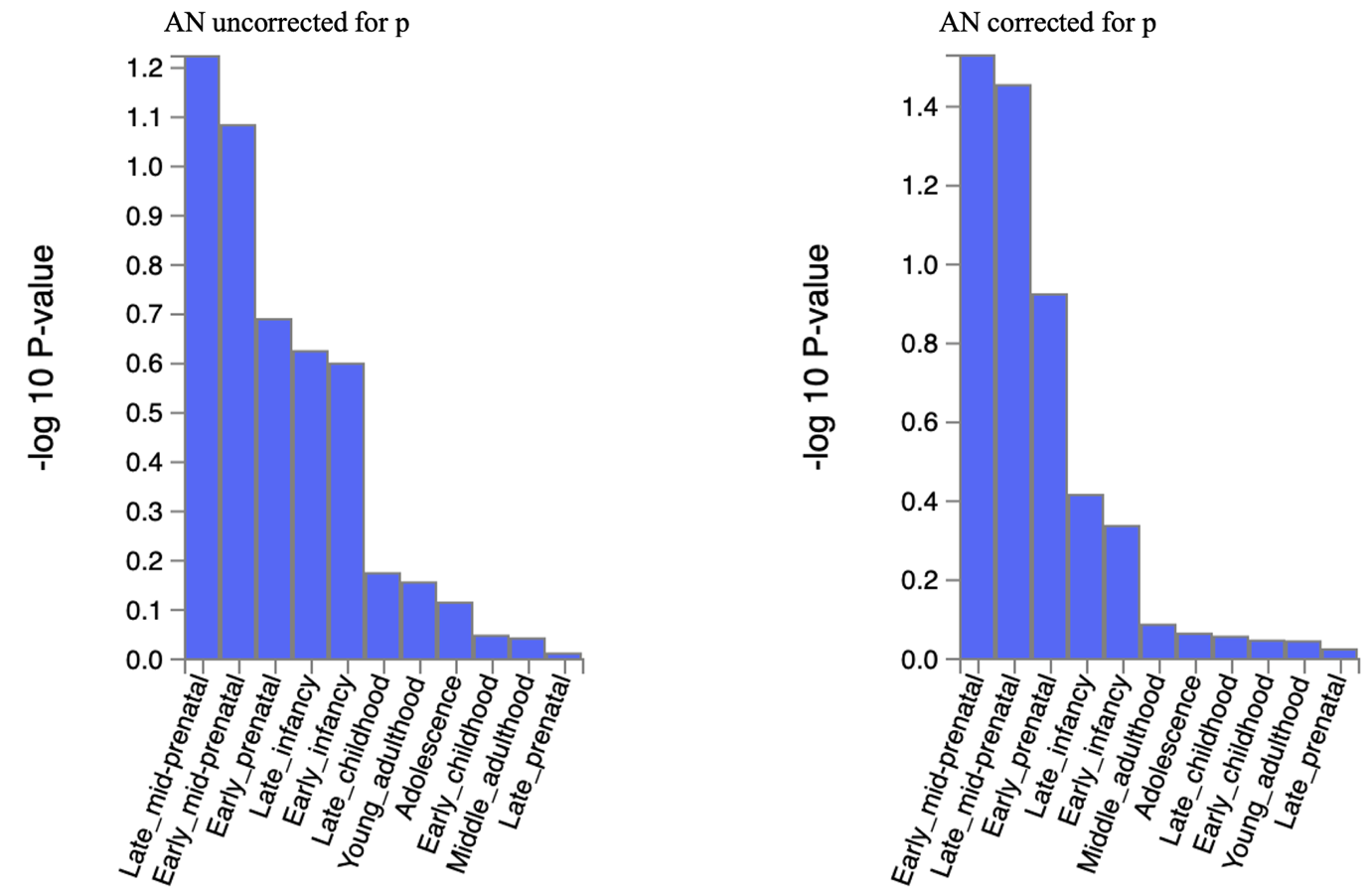

**Supplementary** **Figure 25. The brain sample enrichment results on 11 general brain developmental stages by BrainSpain of AN uncorrected and corrected for p**

Results (-log10 (one-sided P-value)) from MAGMA gene-property analysis of relationships between gene expression data of developmental brain samples (BrainSpan) and AN associations before and after correcting for p. The test was performed for average gene-expression per. brain sample conditioning on the average expression across all developmental stages. The full results are available in Supplementary Tables 30-33.

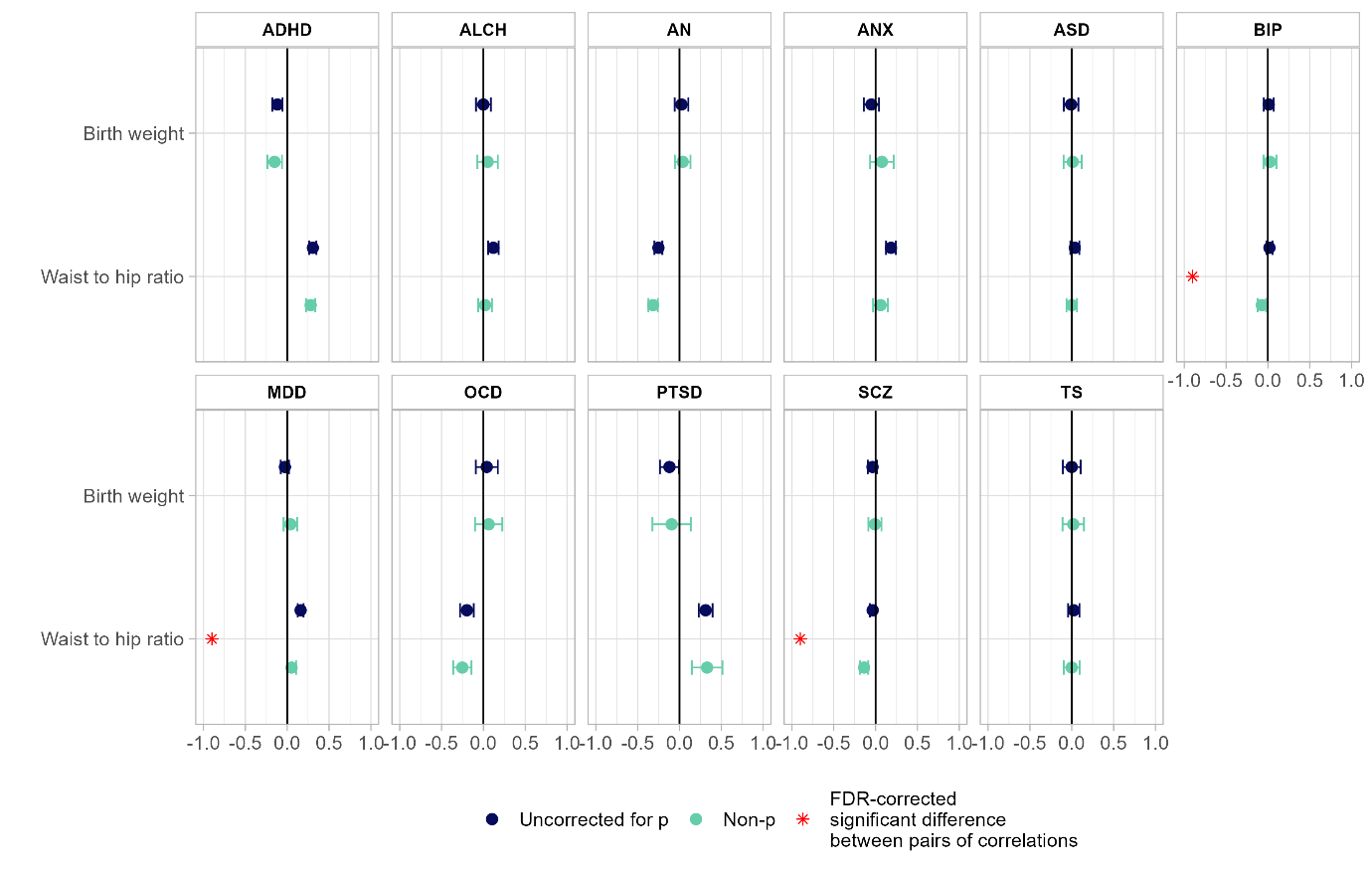

**Supplementary** **Figure 26. Genetic correlations between 11 major psychiatric disorders and anthropometric traits before and after accounting for p.** The dots represent genetic correlations estimated using LDSC regression. Correlations with psychiatric disorders uncorrected for p are in blue, with psychiatric disorders corrected for p in green. Error bars represent 95% confidence intervals. Red asterisks indicate a statistically significant (FDR-corrected *P* < 0.05, two-tailed test) difference in the magnitude of the correlation with disorders uncorrected for p versus disorders corrected for p. Exact *P* values for all associations are reported in Supplementary Table 40.

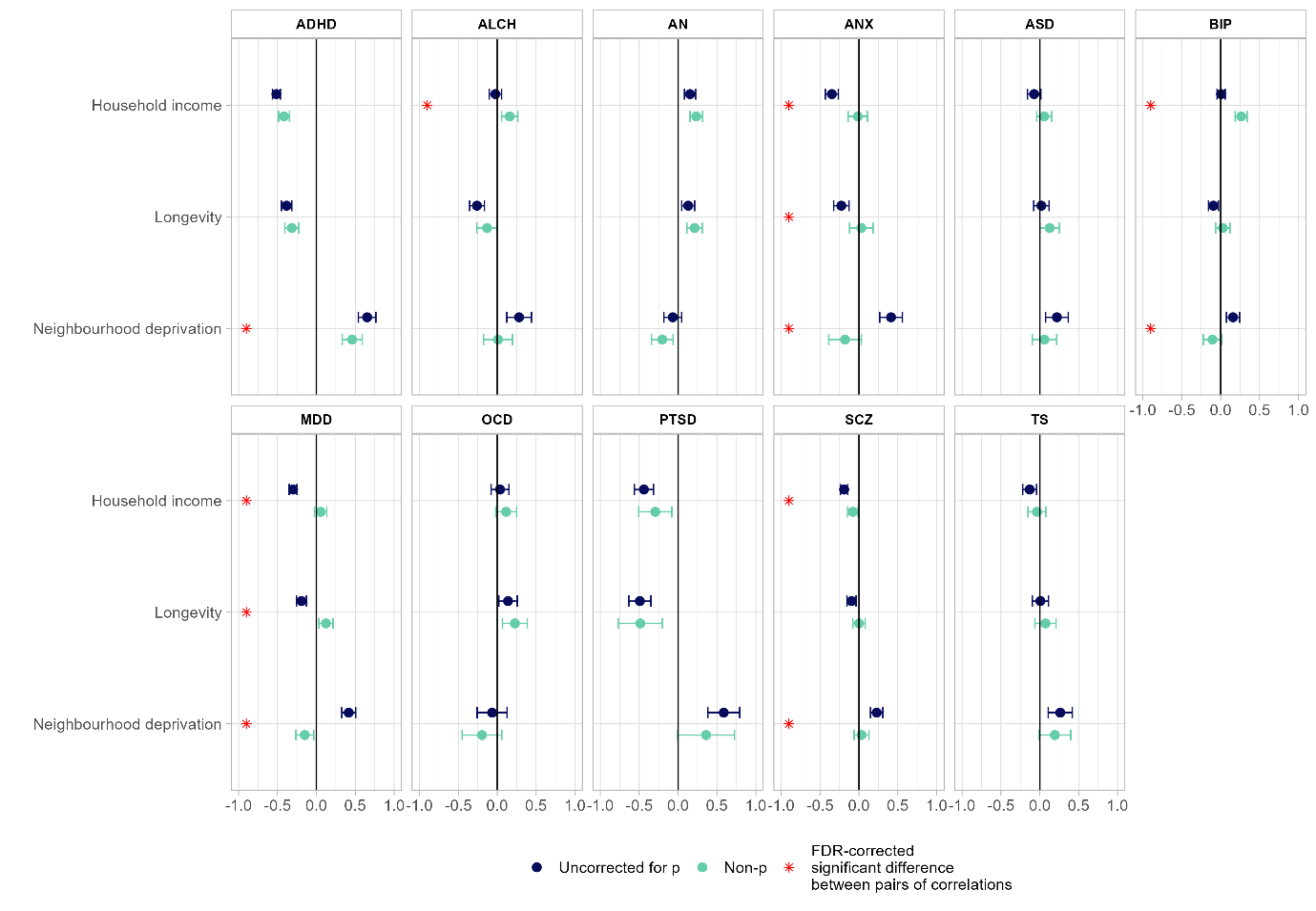

**Supplementary** **Figure 27. Genetic correlations between 11 major psychiatric disorders and socio-demographic traits before and after acounting for p.** The dots represent genetic correlations estimated using LDSC regression. Correlations with psychiatric disorders uncorrected for p are in blue, with psychiatric disorders corrected for p in green. Error bars represent 95% confidence intervals. Red asterisks indicate statistically significant (FDR-corrected *P* < 0.05, two-tailed test) differences in the magnitude of the correlation with disorders uncorrected for p versus disorders corrected for p. Exact *P* values for all associations are reported in Supplementary Table.
